# <u>C</u>N-105 in Participants with <u>A</u>cute Supra<u>T</u>entorial Intra<u>C</u>erebral <u>H</u>emorrhage (CATCH) Trial

**DOI:** 10.1101/2020.10.13.20211417

**Authors:** Michael L. James, Jesse Troy, Nathaniel Nowacki, Jordan Komisarow, Christa B. Swisher, Kristi Tucker, Kevin Hatton, Marc A. Babi, Bradford B. Worrall, Charles Andrews, Daniel Woo, Peter G. Kranz, Christopher Lascola, Maureen Maughan, Daniel T. Laskowitz, for the CATCH Investigators

## Abstract

**Background:** Endogenous apoliloprotein E mediates neuroinflammatory responses and recovery after brain injury. Exogenously administered apolipoprotein E-mimetic peptides can effectively penetrate the brain and down-regulate acute inflammation. CN-105 is a novel apolipoprotein E-mimetic pentapeptide with excellent preclinical evidence as an acute intracerebral hemorrhage (ICH) therapeutic. The <u>C</u>N-105 in participants with <u>A</u>cute supra<u>T</u>entorial intra<u>C</u>erebral <u>H</u>emorrhage (CATCH) trial is a first-in-disease-state, multi-center, open-label trial evaluating safety and feasability of CN-105 administration in patients with acute primary supratentorial ICH.

**Methods:** Eligible patients were age 30-80 years, had confirmed primary supratentorial ICH, and able to intiate CN-105 administration (1.0 mg/kg every 6 hours for 72 hours) within 12 hours of symptom onset. *A priori* defined safety endpoints, including hematoma volume, pharmacokinetics, and 30-day neurological outcomes were analyzed. For comparisons, CATCH participants were matched 1:1 with a contemporary ICH cohort through random selection. Hematoma volumes determined from computed tomography images on Days 0, 1, 2, and 5 and ordinal modified Rankin Score at 30 days after ICH were compared.

**Results:** In 39 participants enrolled across six study sites in the United States, adverse events occurred at expected rate without increase in hematoma expansion or neurological deterioration or significant serum accumulation. CN-105 treatment had an odds ratio (95% confidence interval) of 2.69 (1.31–5.51) for lower 30-day mRS, after adjustment for ICH Score, sex, and race/ethnicity, compared to matched contemporary cohort.

**Conclusion:** CN-105 administration represents an excellent translational candidate as an actue ICH therapeutic due to its safety, dosing feasibility, favorable pharmacokinetics, and evidence of improved neurological recovery.

## Introduction

Spontaneous non-traumatic intracerebral hemorrhage (ICH) accounts for 10-15% of all strokes, with an incidence of approximately 25 per 100,000 worldwide.^1^ ICH is also associated with disproportionate morbidity and mortality as compared to other forms of stroke, and nearly half of afflicted patients will die within 30 days of hemorrhage.^2^ Despite the high personal, societal, and financial impact of ICH, little improvement has been made in ICH-related mortality over the last two decades, and effective pharmacological treatments for ICH remain a compelling unmet need.^3, 4^

Although recent clinical trials have focused on interventions to reduce hematoma extension after ICH via reduction of blood pressure,^5, 6^ manipulation of the coagulation system,^7-9^ and minimaly invasive evacuation techniques,^10^ another viable strategy may be targeting neuroinflammatory responses. The acute CNS response to ICH is characterized by the recruitment of hematogenous inflammatory cells and the activation of endogenous microglia and astrocytes. This process leads to up-regulation of proinflammatory cytokines, production of free radicals, and neuronal excitotoxicity. These neuroinflammatory responses contribute to oxidative injury, breakdown of the blood-brain barrier, and secondary tissue injury.^11^ Thus, reduction of the acute neuroinflammatory response may result in reduced secondary tissue injury and improved outcomes.

Apoliloprotein E (apoE) is a key mediator of the neuroinflammatory response and recovery from brain injury.^12, 13^ Three common human apoE isoforms, designated apoE2, apoE3 and apoE4, differ by single cysteine to arginine substitutions at positions 112 and 158. ApoE3 plays an adaptive role in downregulating glial activation and reducing secondary neuronal injury.^14-16^ Conversely, apoE4 is associated with increased inflammation and poor neurobehavioral outcome.^17, 18^ We have previously demonstrated that small apoE-mimetic peptides can effectively penetrate the brain and down regulate acute neuroinflammatory responses *in vivo*.^13, 14, 19-27^ CN-105 is a small, 5-amino acid apoE-mimetic peptide derived from the polar face of the receptor-binding region of apoE and was optimized to retain the anti-inflammatory and neuroprotective effects of the holoprotein, while retaining good CNS penetration and minimal toxicity.^28^ In a randomized, placebo-controlled phase 1 trial, escalating and repeated doses of CN-105 were found to be safe and well tolerated in healthy adults.^29^

Now, we report the results of a first-in-disease-state, multi-center, open-label trial evaluating safety and feasability of CN-105 administration in patients with acute primary supratentorial ICH. We also report CN-105 effects on hematoma volume, 30-day mortality, and neurological outcomes, including comparisons with matched controls from a large contemporary cohort.

## Methods

The <u>C</u>N-105 in participants with <u>A</u>cute supra<u>T</u>entorial intra<u>C</u>erebral <u>H</u>emorrhage (CATCH) Trial was designed as a multicenter, open label, phase 2 trial of CN-105 in patients with acute primary supratentorial ICH. The primary objective of the study was to assess safety and feasability of CN-105 administration within 12 hours of acute primary ICH. Secondary objectives of the study were to define the pharmacokinetic profile of CN-105 in this patent population, determine target engagement, and evaluate early signals of therapeutic efficacy. CATCH was approved by the central institutional review board Copernicus Group Independent Review Board and by each participating sites’ institutional review board and has been performed in accordance with the ethical standards as laid down in the 1964 Declaration of Helsinki and its later amendments. Informed consent was obtained from all individual participants included in the study.

### Study Population

CATCH enrolled a total of 39 participants across six study sites in the United States. To be eligible for participation, patients had to be age 30-80 years (inclusive), have a radiographically confirmed diagnosis of primary supratentorial ICH, and have CN-105 administration initiated within 12 hours of symptom onset. A full set of inclusion and exclusion criteria may be found in Supplementary Table 1. CN-105 treated participants were compared to a matched cohort of participants drawn from the Ethnic/Racial Variations of Intracerebral Hemorrhage (ERICH) study, a cohort of 3000 patients with ICH prospectively assembled between 2010 and 2015.^30^ CATCH subjects were matched for admission ICH Score,^31^ sex, and race/ethnicity. Supplemental Figure demonstrates the ERICH-CATCH matching process.

**Table 1.** Participant Characteristics. Characteristics of Ethnic/Racial Variations of Intracerebral Hemorrhage (ERICH) study participants matched to CN-105 in Participants with Acute SupraTentorial Intracerebral Hemorrhage (CATCH) Trial participants. Note: %, percentage; _1_, Wilcoxin ranked sums; _2_, chi-square; n, number; SD, standard deviation.

|  | CATCH<br>(N=38) | ERICH<br>(N=240) | p value |
| --- | --- | --- | --- |
| **Age** |  |  | 0.485<sup>1</sup> |
| Median | 62.00 | 59.50 |  |
| Range | (32.00-80.00) | (21.00-85.00) |  |
| **Female Sex** | 15 (39.5%) | 96 (40.0%) | 0.951<sup>2</sup> |
| **Caucasian Race** | 19 (50.0%) | 161 (67.1%) | 0.008<sup>2</sup> |
| **Hispanic Ethnicity** | 0 (0.0%) | 104 (43.3%) | <0.001<sup>2</sup> |
| **ICH Score** |  |  | 0.556<sup>1</sup> |
| 0 | 15 (39.5%) | 79 (32.9%) |  |
| 1 | 11 (28.9%) | 87 (36.3%) |  |
| 2 | 10 (26.3%) | 48 (20.0%) |  |
| 3 | 1 (2.6%) | 20 (8.3%) |  |
| 4 | 1 (2.6%) | 6 (2.5%) |  |
| **BMI at Screening** |  |  | 0.357<sup>1</sup> |
| Median | 27.65 | 29.03 |  |
| Range | (18.79-58.91) | (17.63-72.60) |  |
| **Total Number of Infusions** |  |  |  |
| Mean (SD) | 12.68 (1.21) |  |  |

### Study Procedures

Patients meeting eligibility criteria for the trial were approached for consent. After consent was signed by the patient or their legal authorized representative, participants received intravenous CN-105 administered over 30 minutes, at a dose of 1.0 mg/kg every six hours for up to a maximum of 13 doses (72 hours) or until discharge. Participants were monitored daily throughout the treatment phase of the study (up to a maximum of five days) and received standard of care as determined by their attending physician for the duration of the study. The ICH score^31^ was calculated for each patient at screening. Severity of neurological deficit was assessed using the National Institutes of Health Stroke Scale Score (NIHSS) and Glasgow Coma Score (GCS) on presentation, and daily for 5 days after ICH onset, death or discharge. After discharge from the hospital, participants entered a three-month follow up phase with in-person clinic visit at 30 days and a follow-up telephone interview at 90 days.

### Safety Endpoints

Adverse events (AE) were recorded starting immediately after obtaining informed consent and extending through the last study-related procedure completed up to Day 90 follow up phone call. At each study visit, the study teams queried about AEs and Serious AEs (SAE) since the last study visit. Events were followed for outcome information until resolution, stabilization, or end of study. *A priori* defined safety endpoints included: 1) number and severity of AEs/SAEs throughout study duration; 2) in-hospital 30- and 90-day mortality; 3) treatment-related mortality; 4) in-hospital neurological deterioration, defined as an increase of NIHSS>2 from baseline, persisting more than 24 hours and unrelated to sedation; 5) incidence of cerebritis, meningitis, ventriculitis; 6) incidence of systemic infection; and 7) incidence of hematoma expansion >30% from baseline computed tomography (CT) head imaging.

### Pharmacokinetic Analyses

A subpopulation of 7 participants receiving CN-105 underwent serial blood sampling for noncompartmental pharmacokinetic (PK) profiling. PK samples were collected prior to dosing (within 1 hour), 0.083, 0.167, 0.5, 1, 2, 4, 5.5 hours after start of each dose for the first 2 doses, prior to dosing (within 1 hour) for each of doses 3 to 13 and 0.083, 0.167, 0.5, 1, 2, 4, 6, 12, 24 hours after the last dose (dose 13). Plasma CN-105 concentrations were determined by MPI Research (Mattawan, Michigan). Plasma samples were stored in 1% HALT with K2EDTA at – 70°C before analysis. Liquid chromatography-tandem mass spectrometry analysis was performed using a positive Turbo IonSpray® interface on a Sciex API-5000 (Applied Biosystems, Foster City, California) and multiple reaction monitoring. The analytical range was 1.00 ng/mL to 1000 ng/mL. The assay precision in quality-control samples was <20% for the lowest limit of quantitation and < 15% for all other concentrations.

### Imaging Outcomes

All participants underwent initial diagnostic CT scans for confirmation of ICH and presence of any underlying structural or vasular abnormality which would preclude eligibility. All participants underwent follow-up CT scans were performed at 24 hours (Day 1), 48 hours (Day 2) and 120 hours (Day 5) after initial diagnostic CT. For each CT scan, hematoma location, volume, and intraventricular extension were determined. Hematoma volumes were measured using previously described techniques for semiautomated segmentation on commercially available software (AnalyzePro v 1.0; AnanlyzeDirect, Inc., Chicago, IL).^32^ Briefly, hematoma boundaries were established by using an edge-detection tool using an attenuation threshhold range of 42-100 Hounsfield Units. Intial segmentation by the tool was reviewed and adjusted by study neuroradiologist to ensure accuracy before hematoma volumes were automatically calculated from the segmented images.

### Neurological Outcomes

Investigation-related clinical examinations were performed at screening and then daily for the first 5 days or until hospital discharge, whichever came first. After discharge, in-person clinic visit was performed at 30 days and a telephone interview at 90 days after enrollment. Modified Rankin Scale (mRS), NIHSS, Stroke Impact Scale-16, Barthel Index and Montreal Cognitive Assessment were used to assess CN-105 effects on neurological and cognitive function. Comparison of 30-day mRS between participants treated with CN-105 and matched ERICH controls was the primary analysis of efficacy.

### Statistical Approach

All participants who received at least one dose of CN-105 were included in analysis of safety outcomes using descriptive methods. For comparison with the ERICH study, CATCH and ERICH participants were matched 1:1 based on initial ICH score and sex through random selection from a sampling frame of 240 eligible controls (Supplemental Figure). Hematoma volumes determined from CT images on Days 0, 1, 2, and 5 were compared descriptively to matched ERICH participants CT images on Days 0-5 after ICH using box plots. Noncompartmental PK analysis was performed using the Phoenix WinNonlin (version 6.3) software. Analysis of mRS was based on rational imputation as follows: three participants were lost to follow-up after discharge from the hospital to home (two were lost to follow-up at Days 30 and 90 and one at Day 90 only). These missing values were imputed with their last observed value, given their discharge to home (mRS=2 for 2 participants and mRS=1 for one participant). Three participants had interior missing data at Day 30 which was imputed as their Day 5 observed value (mRS=4 for all 3 participants). Distribution of Day 30 mRS was compared between CATCH and matched controls from the ERICH study using a proportional odds model with the robust (sandwich) variance estimator. Additional sensitivity analyses were conducted to evaluate results after adjustment for race and age. Odds ratios and 95% confidence intervals are reported from the proportional odds model. All statistical tests were 2-sided and used alpha=0.05. Analyses were conducted in SAS 9.4 (SAS Institute, Cary, NC).

## Results

Between August 2017 and March 2019, 39 patients with acute ICH at six participating medical centers were consented. One patient was excluded after consent due to ICH secondary to aneurysm; all 38 participants enrolled into the study received CN-105 with 35 participants receiving their first dose less than 12 hours from ICH onset,10 first doses occurring in under 6 hours, and 23 participants completing all 3 days of dosing. CATCH participants were matched 1:1 to ERICH participants. Characteristics of the study and comparator cohorts are summarized in Table 1.

Non-serious AEs are summarized in Table 2. No significant safety concerns were identified with CN-105 administration in participants with ICH. Throughout the study, vital signs were stable and did not change significantly. No participant experienced changes in electrocardiogram parameters. Clinical chemistry, blood counts, coagulation profiles, and lipid parameters were stable and did not change significantly throughout the course of the study. No participants experienced central nervous system (CNS) infection. Fourteen nonfatal SAEs were experienced by 12 (32%) participants, reported most commonly in the Nervous System and Respiratory Disorders with 4 events in each category. All SAEs are listed in Supplemental Table 2. Thirty-one (82%) participants experienced a treatment emergent AE, summarized in Table 2. The most commonly experienced treatment emergent AEs (≥15% of participants) were pain (29%), constipation (26%), leukocytosis (24%), hypophosphatemia (21%), hypomagnesemia (18%), hyponatremia (18%), nausea (18%), and pyrexia (16%). Non-serious treatment emergent AEs are listed in Supplemental Table 3. Eleven treatment emergent SAE’s were experienced by 9 (24%) participants (Supplemental Table 4), ultimately deemed unlikely related to study drug. Five (13%) participants experienced in-hospital neurological deterioration during the course of the study. Nine (24%) participants had incidence of hemorrhage extension throughout the study, one with clinical deterioration. Mean (SD) hematoma volumes by CT were 22.8 (23.7), 25.1 (25.1), 21.9 (24.9), and 19.7 (19.6) cm^3^ on admission, Day 1, Day 2, and Day 5 after ICH, respectively. These volumes were comparable to hematoma volumes in matched ERICH participants with ICH (Figure 1). Two (5%) participants received treatment for elevated intracranial pressure; both participants died in-hospital due to brain herniation from the index ICH. Three (8%) participants died prior to 30-day follow up. All 3 participants experienced in-hospital mortality due to adverse events related to the primary disease and within 6 days of the first dose of CN-105.

**Table 2.** Non-Serious AE Summary. Summary of all non-serious adverse events (AEs). Note: #, number; ^a^, The number of subjects with mild, moderate, and severe events may sum to more than the total number of subjects because some subjects experienced more than one event.

|  | All Non-Serious AEs | Treatment Emergent Non-Serious AEs | Related, Treatment Emergent Non-Serious AEs |
| --- | --- | --- | --- |
| **# of Events** | 237 | 230 | 86 |
| Mild | 183 | 177 | 73 |
| Moderate | 51 | 50 | 13 |
| Severe | 3 | 3 | 0 |
| **# of Subjects<sup>a</sup>** | 31 | 30 | 16 |
| Mild | 30 | 29 | 16 |
| Moderate | 19 | 19 | 5 |
| Severe | 2 | 2 | 0 |

**Table 3.**
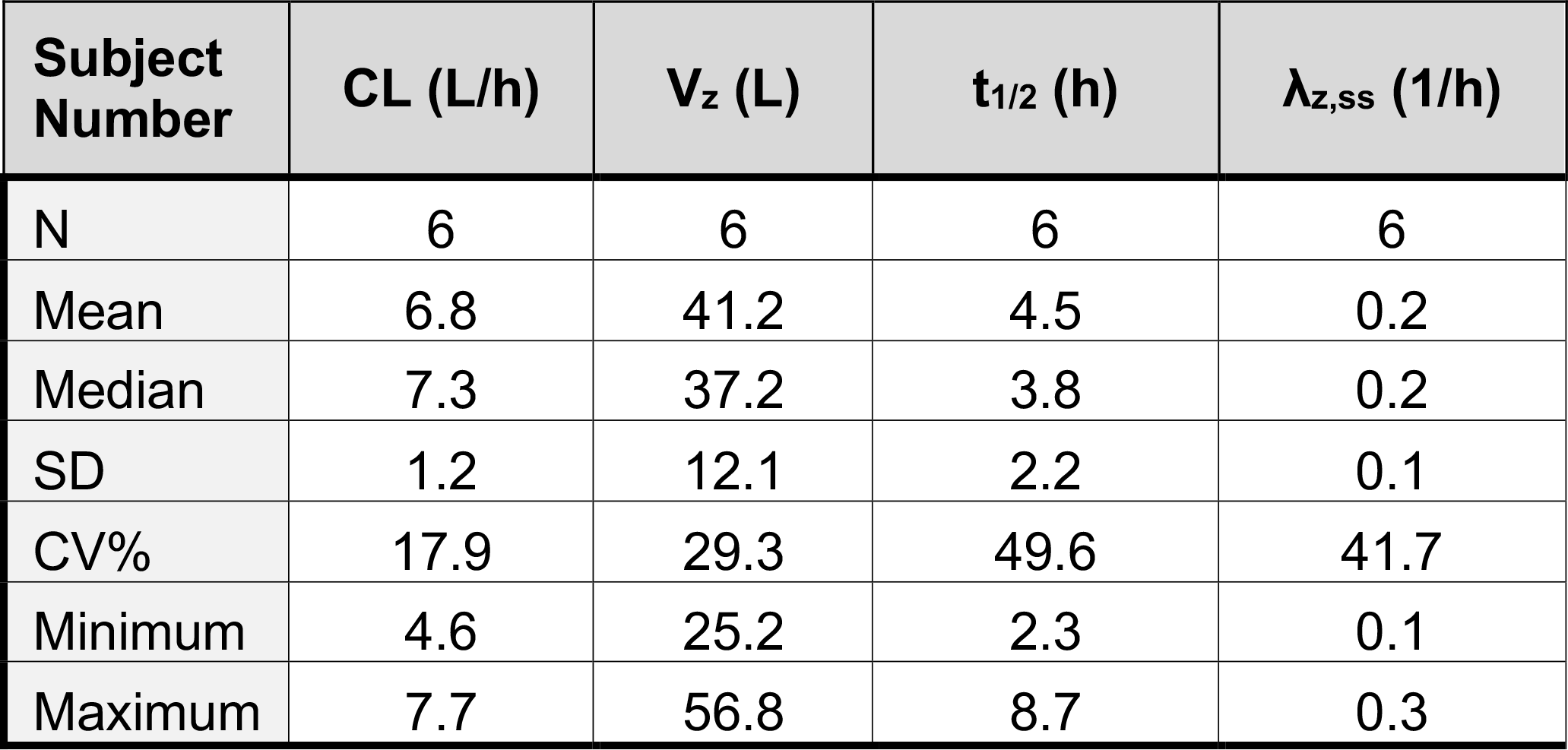
CN-105 PK parameters. Pharmacokinetic profile of CN-105 from a subpopulation of 7 participants in the CN-105 in Participants with Acute SupraTentorial Intracerebral Hemorrhage (CATCH) Trial. Note: %, percentage, λ_z,ss_, Terminal elimination rate constant after last dose; CL, clearance; h, hours; L, liter; N, number; SD, standard deviation; t_1/2_, terminal phase disposition half-life; V_z_, volume of distribution during terminal phase.

| **Subject Number** | **CL (L/h)** | **V<sub>z</sub> (L)** | **t<sub>1/2</sub> (h)** | **λ<sub>z,ss</sub> (1/h)** |
| --- | --- | --- | --- | --- |
| N | 6 | 6 | 6 | 6 |
| Mean | 6.8 | 41.2 | 4.5 | 0.2 |
| Median | 7.3 | 37.2 | 3.8 | 0.2 |
| SD | 1.2 | 12.1 | 2.2 | 0.1 |
| CV% | 17.9 | 29.3 | 49.6 | 41.7 |
| Minimum | 4.6 | 25.2 | 2.3 | 0.1 |
| Maximum | 7.7 | 56.8 | 8.7 | 0.3 |

**Figure 1.**
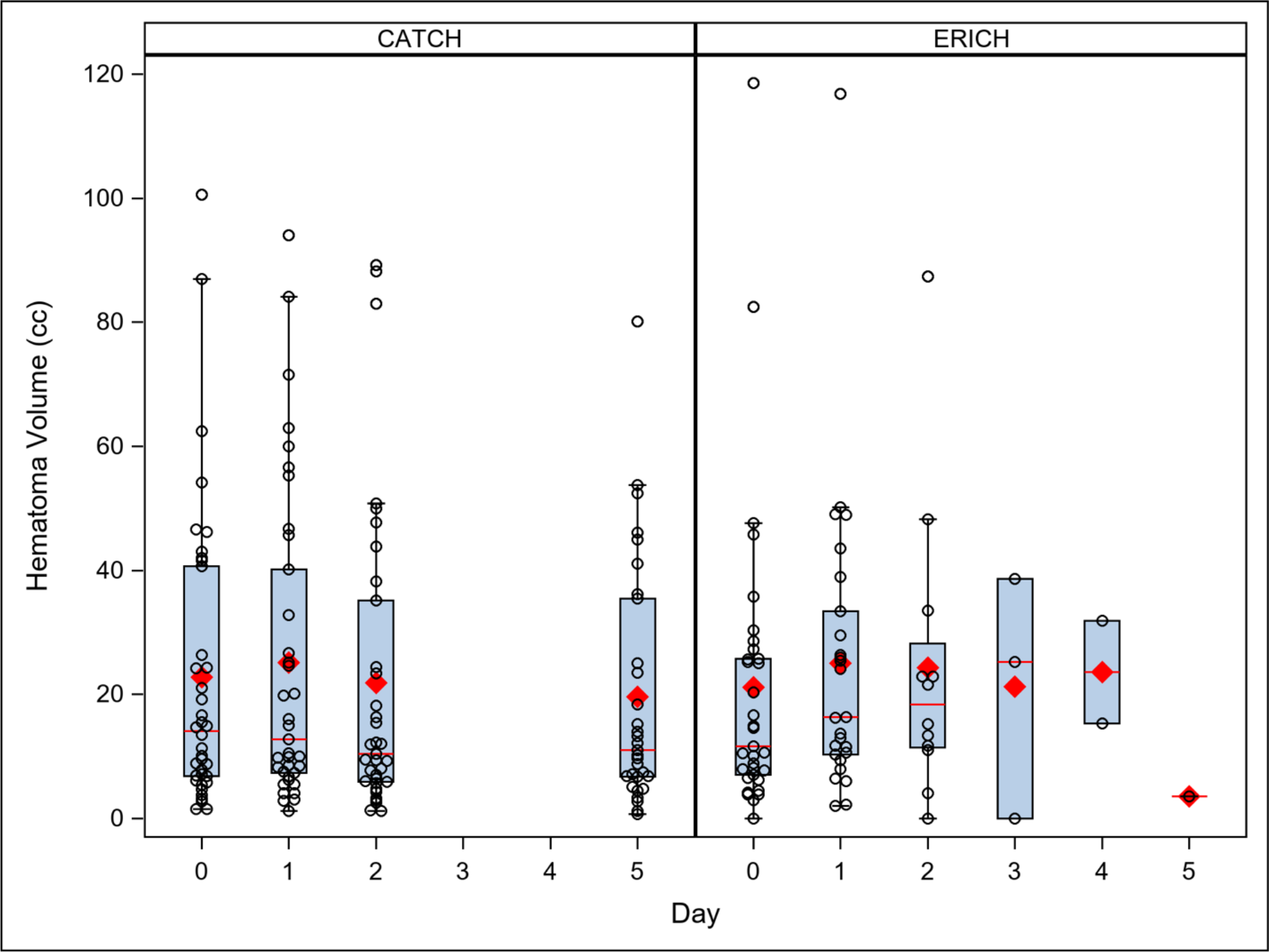
Hematoma volume comparison between CATCH and matched ERICH participants. Comparison of hematoma volumes in Ethnic/Racial Variations of Intracerebral Hemorrhage (ERICH) study participants matched to CN-105 in Participants with Acute SupraTentorial Intracerebral Hemorrhage (CATCH) Trial participants. Hematoma volumes volumetrically measured from brain computed tomography images on Days 0, 1, 2, and 5 after enrollment from 38 CATCH participants were compared descriptively to 1:1 matched ERICH participants on Days 0-5 after intracerebral hemorrhage using box plots. No difference in hematoma volumes were noted. Note: cc, cubic centimeters.

As shown in Figure 2, trough plasma concentrations for all subjects after Dose 1 demonstrated comparable exposures without evidence of clinically significant accumulation. Mean (SD) maximum concentrations were 5610.0 ng/mL (1335.1), 6296.7 ng/mL (2155.8), and 5638.3 ng/mL (1335.9) for dose 1, 2, and 13, respectively. The mean (SD) area under the curve (AUC) were 11206.2 ng*hr/mL (3863.4), 12936.4 ng*hr/mL (4983.6), 13347.0 ng*hr/mL (3369.1) for dose 1, 2, and 13, respectively. Accumulation of drug during multiple dosing was minimal with mean (SD) ratios of 1.2 (0.1) and 1.3 (0.4) of Dose 1:Dose 2 and Dose 1:Dose 13 AUCs, respectively. Starting from Dose 4, trough concentrations after each dose were not significantly different from the subsequent doses, through Dose 13. Table 3 demonstrates PK parameters of CN-105 after acute ICH. Plasma concentrations of CN-105 in one participant were less than 4% of mean concentrations of other participants from 5 minutes after the start of of the initial infusion to 0.5 hours after the stop of infusion. Because of the grossly abnormal PK profile in this participant after Dose 1, the paticipant’s measurements were censored.

**Figure 2.**
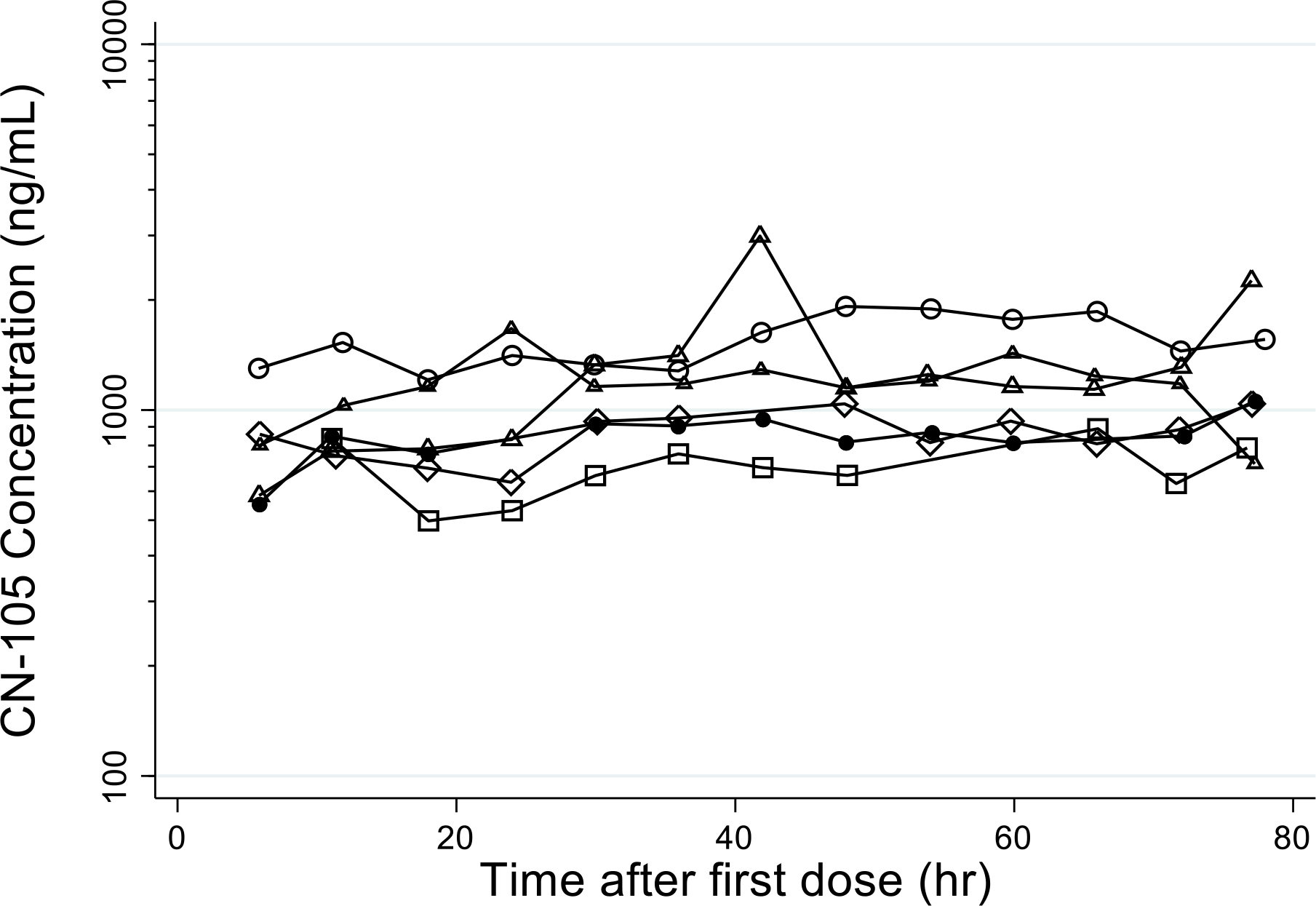
Pharmacokinetics of CN-105 in participants with ICH. Plot of trough plasma concentrations after the first dose of CN-105 from a subpopulation of 7 participants in the CN-105 in Participants with Acute SupraTentorial Intracerebral Hemorrhage (CATCH) Trial demonstrated comparable exposures without evidence of clinically significant accumulation. Note: hr, hour; ml, milliter; ng, nanogram.

*A priori* primary neurological outcome was mRS from in-person 30 day assessment. Compared to matched participants in the ERICH study, CATCH trial participants had significantly improved 30-day mRS outcomes (Figure 3). Secondary analyses of mRS, NIHSS, Stroke Impact Scale-16, Barthel Index, and Montreal Cognitive Assessment at hospital discharge and 30 days after ICH are summarized in Supplemental Table 5; however, the ERICH study lacked appropriate comparator data for these outcomes. Over the course of CN-105 treatment, NIHSS improved from median (range) of 12.0 (4.0, 25.0) to 8.5 (1.0, 29.0). Similarly, mRS in CATCH participants improved from Day 5 through Day 90 (Figure 3).

**Figure 3.**
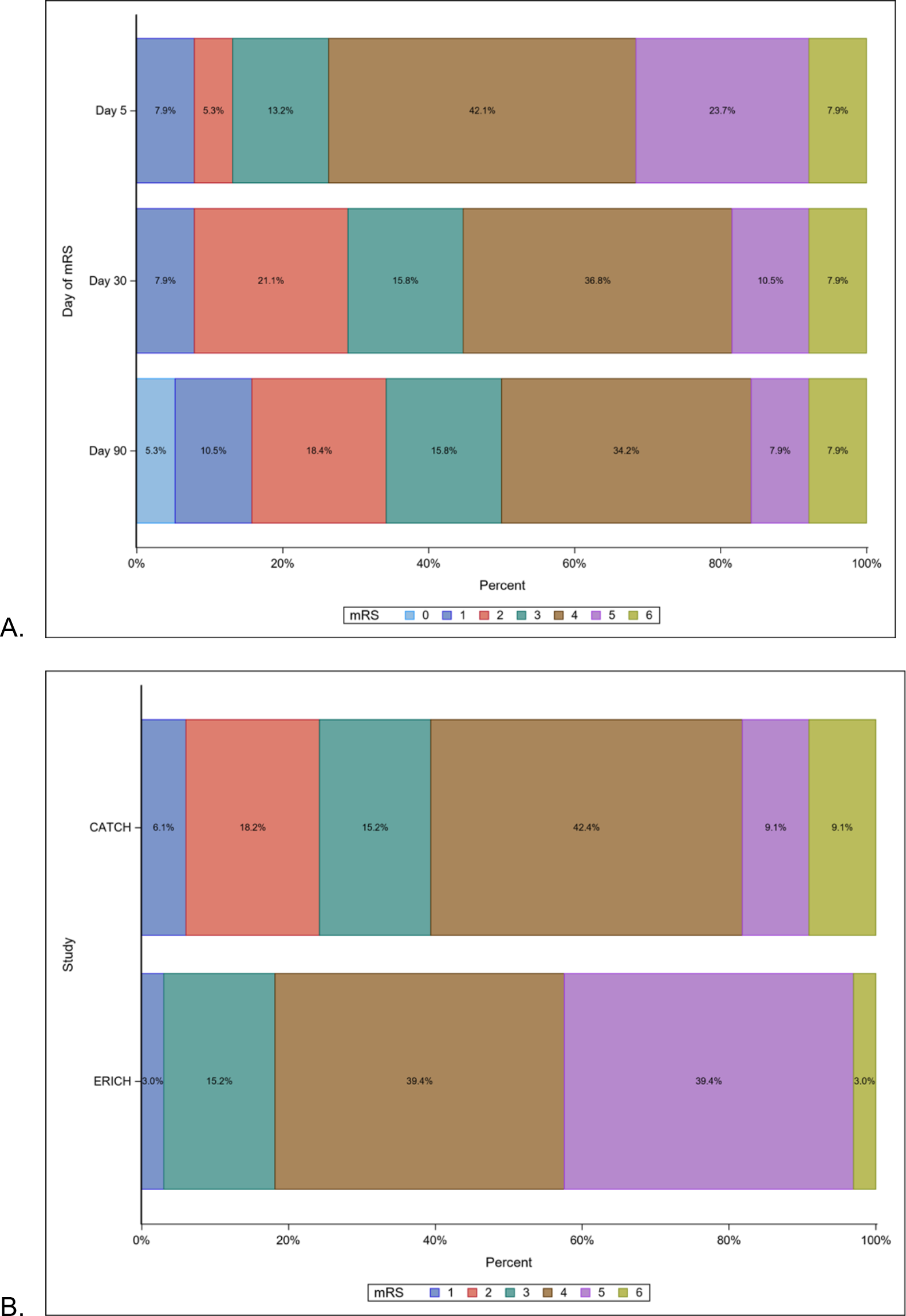
CATCH participants mRS at 5-, 30-, & 90-days after ICH (A) and 30-day mRS comparison between CATCH and matched ERICH participants (B). Modified Rankin Scale (mRS) from CN-105 Participants with Acute SupraTentorial Intracerebral Hemorrhage (CATCH) Trial and matched Ethnic/Racial Variations of Intracerebral Hemorrhage (ERICH) study participants. (A) mRS from 38 CATCH trial participants at Days 5, 30 and 90 after enrollment found common odds ratio (95% confidence interval) of 1.63 (1.23 - 2.15) for lower mRS as participants progress from 5 days to 90 days after intracerebral hemorrhage. (B) CATCH trial participants compared to 1:1 matched participants in the ERICH study (n=33) had odds ratio (95% confidence interval) of 2.69 (1.31 – 5.51) for lower 30-day mRS, after adjustment for ICH Score, sex, and race/ethnicity.

## Discussion

Administration of 1.0 mg/kg CN-105 every 6 hours for 72 hours is safe and feasible when intiated within 12 hours of spontaneous ICH. In this critically ill population of patients with acute ICH, SAEs and AEs occurred at a rate expected from our other clinical trials. Further, CN-105 was not associated with an increase in hematoma expansion or unanticipated neurological deterioration. CN-105 PK in this cohort of participants with ICH was similar to the previously reported PK in healthy subjects, without significant accumulation and reaching steady state after the fourth dose. Finally, treatment with CN-105 was associated with improved 30-day neurological outcomes as compared to contemporary cohort of closely matched participants from the ERICH database.

Development of the novel apoE-mimetic pentapeptide, CN-105, was based on initial observations that apoE exerted anti-inflammatory and neuroprotective effects in the CNS.^33, 34^ Based on evidence suggesting that these adaptive responses were mediated via interactions with the LRP-1 receptor,^16, 35-37^ a series of apoE-mimetic peptides with therapeutic potential were rationally developed from the apoE receptor binding region.^38-41^ These peptides improved functional outcomes in a wide variety of preclinical cerebrovascular and traumatic brain injury models.^42^ CN-105 was developed from the polar face of the apoE-receptor binding region, and has advantages of increased CNS permeability, potency, and ease of manufacturing as compared to prior apoE-mimetic peptides, which were larger and often contained non-naturally occuring amino acids.^41^ Although the exact mechanism(s) by which CN-105 exerts its protective effects remain incompletely defined, CN-105 improves neurobehavioral outcomes across a variety of preclinical models of acute CNS injuries, including ICH, subarachnoid hemorrhage, ischemic stroke, and traumatic brain injury.^43-46^

APOE genotype correlates with hematoma volume after human ICH.^47, 48^ Thus, effects of CN-105 on hematoma expansion and volume after acute human ICH was a critical *a priori* safety concern. Fortunately, exogenous administration of apoE-mimetic therapies in experimental ICH models has not resulted in greater hematoma volumes or expansion.^14, 22, 24, 44^ In the present study, hematoma volume was unaffected by treatment with CN-105. In fact, injury modifying effects of CN-105 are unlikely to be related to hematoma formation, expansion, or reduction, based on preclincal findings. Rather, evidence suggests that CN-105 reduces brain injury by modifying glial activation and resultant cerebral edema.^43-46^ Since rate of brain edema formation and degree of edema generated has been associated with ICH outcome, CN-105 represents an excellent therapeutic strategy for reducing secondary tissue injury after ICH.^32^ Ongoing work to analyze and compare serum biomarkers of CNS injury and inflammation, as well as perihematomal edema imaging, from participants treated with CN-105 to those not receiving CN-105 is currently underway. Optimization of planned, future Phase 2/3 trials will incorporate these radiographic and biochemical surrogates.

Given its safety and efficacy across a wide variety of preclinical brain injury models, CN-105 is a logical, favorable candidate for clinical translation. However, a large number of promising neuroprotective therapies have failed to successfully translate to improved clinical outcomes,^49^ often due to inadequate preclinical modelling.^3^ To increase probability of successful translation, extensive preclinical studies were performed to demonstrate therapeutic effect of CN-105 in various models of ICH across age, species, and sex (Wang et al., 2020 Neurocrit Care; under review). Further, hypertension is a common comorbidity in patients with ICH (e.g., in this study 21/38 participants were hypertensive, based on blood pressure measurements of systolic ≥140 mmHg or diastolic ≥90 mmHg at screening). Therefore, therapeutic efficacy of CN-105 in the setting of hypertensive comorbidity has also been successfully adressed in preclinical studies (Wang et al., 2020, Neurocrit Care; under review).

Several limitations in the current study should be adressed. Although participants with ICH dosed with CN-105 clearly demonstrated improvement in mRS over time, absence of randomized, contemporaneous placebo-controlled comparator group does not definitively allow us to separate the natural history of recovery from the effect of an investigational drug. However, 30-day mRS of CN-105 treated participants were significantly improved relative to a carefully matched, contemporary cohort of participants drawn from the robustly phenotyped ERICH database, which suggests therapeutic benefit. Unfortunately, comparison for other secondary outcome measures, including NIHSS, Stroke Impact Scale-16, Barthel Index, and Montreal Cognitive Assessment was not possible using the matched cohort from ERICH. However, comparison of these measures against other published clinical trial cohorts do suggest that CN-105 administration may improve neurocognitive outcomes after ICH to a greater degree than might be otherwise expected.^7, 10, 50-52^ Thus, given positive results in clinically relevant ICH preclinical paradigms across species, sex, age, and comorbid hypertension^44^ (Wang et al., 2020 Neurocrit Care; under review), predictable and linear PK profile with favorable safety signal found in prior Phase 1 trial^29^ and the current study, and encouraging safety profile in patients with ICH, CN-105 represents an attractive candidate for clinical translation.

## Conclusion

In this critically ill population of patients with acute ICH, treatment with 1.0 mg/kg of CN-105 administered every six hours for up to three days is safe and feasible with no evidence of drug-related hematoma expansion or neurological deterioration. Treatment with CN-105 was associated with improved mRS at 30 days after ICH when compared to the natural history of a closely matched, contemporary cohort. Due to the open label design, absence of placebo comparator group, or long term functional endpoints, the present results should be interpreted with caution. However, given the wealth of supportive precinical data, favorable PK profile, and clinical safety, CN-105 represents a promising candidate for translation to more definitive efficacy trials in acute ICH incorporating surrogate serological and brain imaging endpoints.

## Data Availability

The data that support the findings of this study are available from the corresponding author upon reasonable request.

## Acknowledgements

The authors would like to express their sincere gratitude to the full list of CATCH Investigators (Supplemental Table 1) for their tireless efforts in executing this trial.

---

**Supplemental Table 1.**
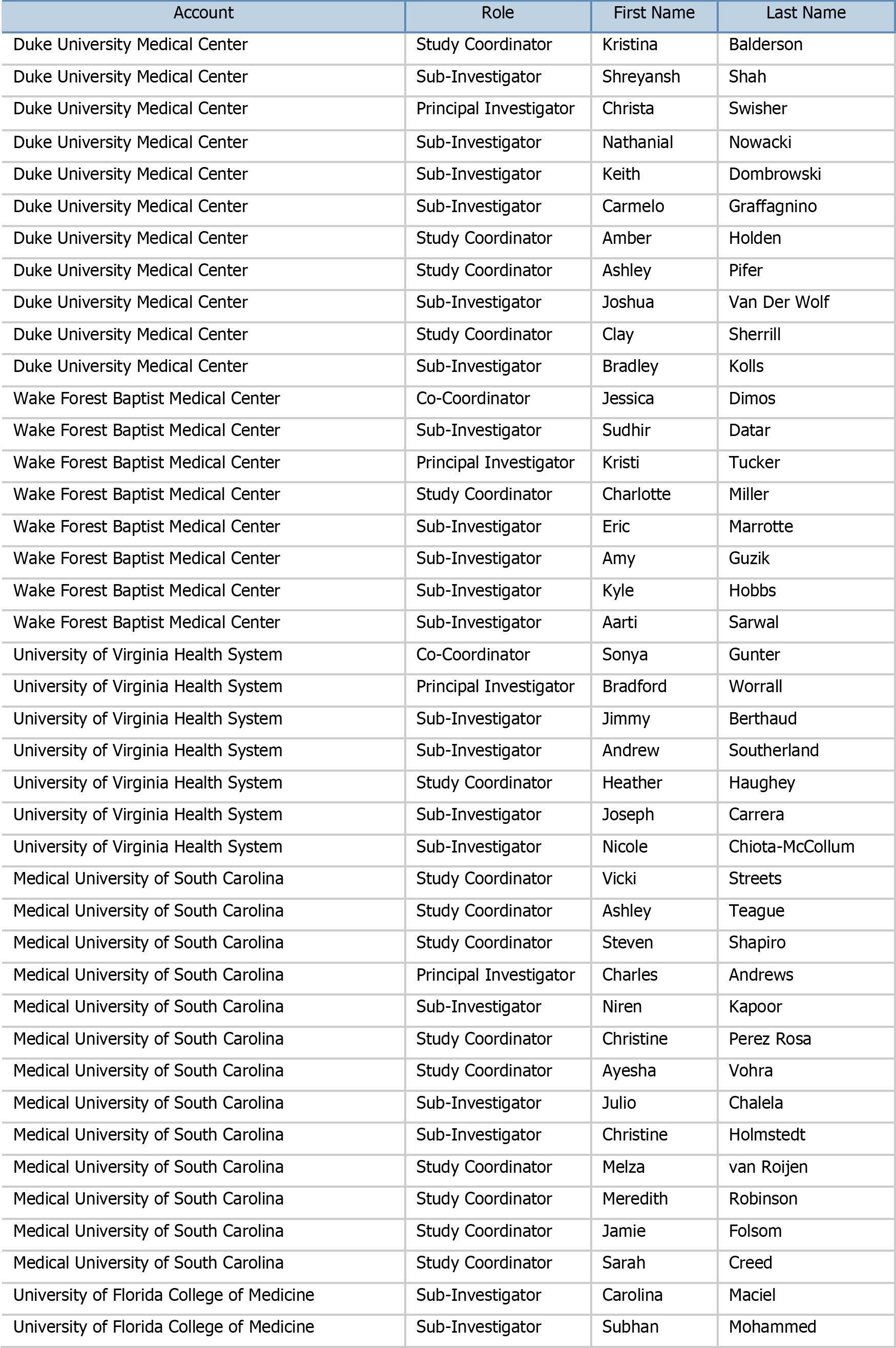
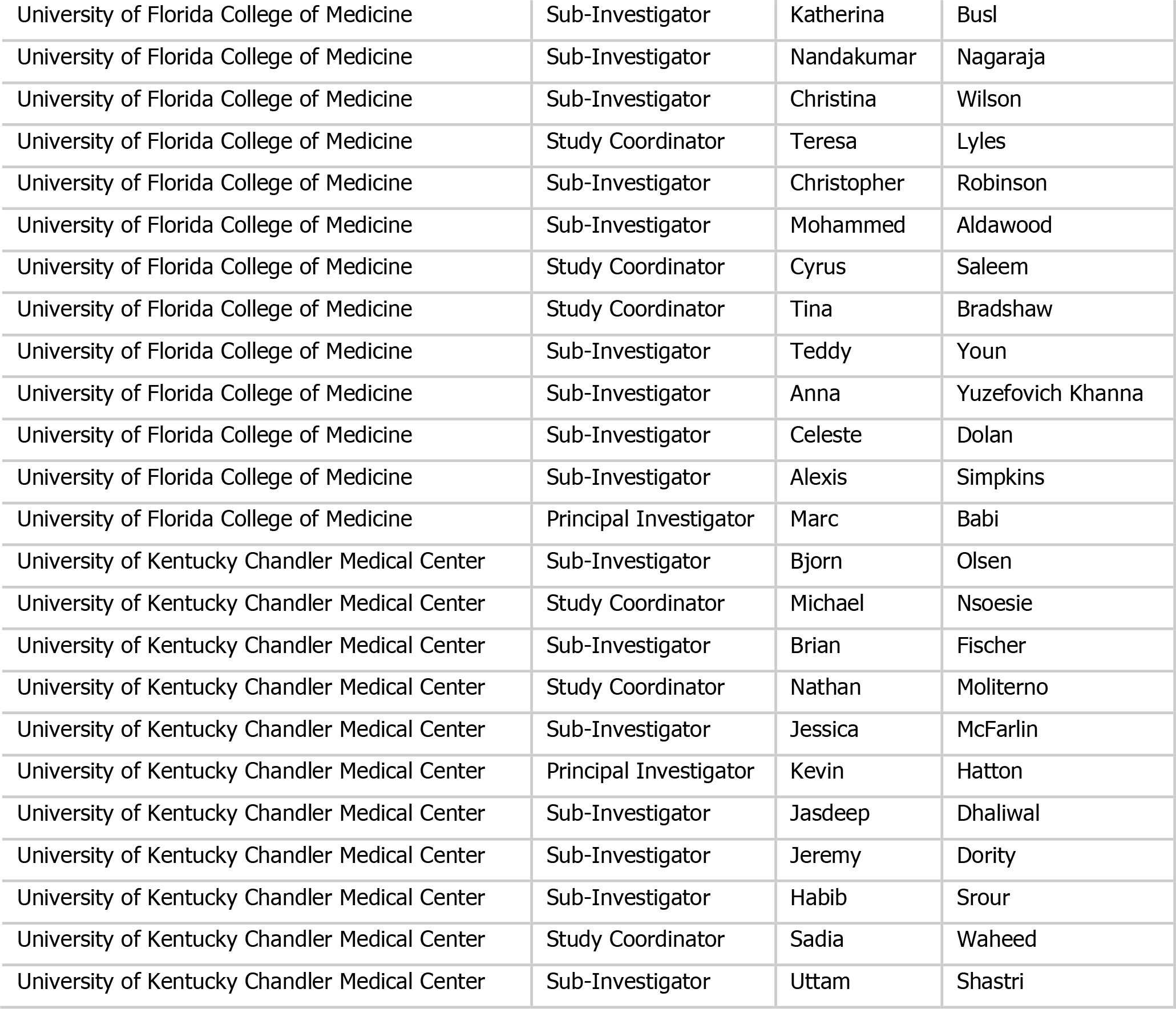
List of CATCH trial Investigators. Full set of inclusion and exclusion criteria for CN-105 in Participants with Acute SupraTentorial Intracerebral Hemorrhage (CATCH) Trial.

**Supplemental Table 2.** Inclusion and exclusion criteria. Full list of all serious adverse events in CN-105 in Participants with Acute SupraTentorial Intracerebral Hemorrhage (CATCH) Trial. Inclusion Criteria:

1. Had given written informed consent to participate in the study in accordance with required regulations; if a participant was not capable of providing informed consent, written consent must be obtained from the participant’s legally authorized representative.
2. Stated willingness to comply with all study procedures and availability for the duration of the study.
3. Was male or female, age 30 to 80 years, inclusive.
4. Had a confirmed diagnosis of spontaneous supratentorial ICH.
5. Able to receive first dose of study drug ≤12 hours after onset of ICH symptoms, such as alteration in level of consciousness, severe headache, nausea, vomiting, seizure, and/or focal neurological deficits, or last known well time.
6. Had an interpretable and measurable diagnostic CT scan.
7. Had a GCS score ≥ 5 on presentation
8. Had a National Institutes of Health Stroke Scale (NIHSS) score ≥ 4
9. Had systolic blood pressure< 200mmHg at enrollment. Exclusion Criteria:

1. Known pregnancy and lactation.
2. Has a temperature greater than 38.5°C at screening.
3. ICH known to result from trauma.
4. Evidence of infratentorial hemorrhage (any involvement of the midbrain or lower brainstem as demonstrated by radiograph or complete third nerve palsy) severely limiting the recovery potential of the patient in the opinion of the investigator.
5. Evidence of primary intraventricular hemorrhage deemed to be at high risk for obstructive hydrocephalus, in the opinion of the investigator or evidence of extra-axial (ie, subarachnoid or subdural) extension of hemorrhage severely limiting the recovery potential of thepatient in the opinion of the investigator.
6. Radiographic evidence of underlying tumor.
7. Known unstable mass or active radiographic evidence and symptoms of herniation syndromes severely limiting the recovery potential of the patient in the opinion of the investigator.
8. Known ruptured aneurysm, arteriovenous malformation, or vascular anomaly.
9. Had a platelet count < 100,000/mL.
10. Had an international normalized ratio> 1.5 or irreversible coagulopathy either due to medical condition or detected before screening.
11. Was taking new oral anticoagulants or low molecular weight heparin at the time of ICH onset.
12. In the opinion of the investigator was unstable and would have benefited from supportive care rather than supportive care plus CN-105.
13. In the opinion of the investigator had any contraindication to the planned study assessments, including CT and MRI.
14. Any condition which could interfere with, or the treatment for which might interfere with, the conduct of the study or which, in the opinion of the investigator, unacceptably increases the individual’s risk by participating in the study.
15. Concomitant enrollment in another interventional study.

**Supplemental Table 3.** Full list of all serious adverse events in CN-105 in Participants with Acute SupraTentorial Intracerebral Hemorrhage (CATCH) Trial.

| Subject ID | Reported Event | MedDRA SOC | MedDRA PT | Relatedness | Severity | Start Day | End Day | Outcome |
| --- | --- | --- | --- | --- | --- | --- | --- | --- |
| 101-003 | Medication administration error | Injury, poisoning and procedural complications | Drug administration error | Unrelated | Moderate | 2 | 2 | Recovered / Resolved |
| 101-004 | Left upper lobe pneumonia | Infections and infestations | Pneumonia | Related | Moderate | 1 | 8 | Recovered / Resolved |
| 101-004 | Failure to thrive | Metabolism and nutrition disorders | Failure to thrive | Unrelated | Moderate | 23 | 57 | Recovered / Resolved |
| 101-004 | Left Frontal IPH | Nervous system disorders | Cerebral haemorrhage | Related | Severe | 58 | 62 | Recovered / Resolved |
| 101-007 | Cerebral herniation | Injury, poisoning and procedural complications | Brain herniation | Unrelated | Severe | 0 | 5 | Fatal |
| 101-008 | Hypoxic Respiratory Failure | Respiratory, thoracic and mediastinal disorders | Respiratory failure | Related | Moderate | 2 | 4 | Recovered / Resolved |
| 101-013 | Urinary Tract Infection (UTI) | Infections and infestations | Urinary tract infection | Unrelated | Moderate | 39 | 44 | Recovered / Resolved |
| 101-015 | Pulmonary Embolism | Respiratory, thoracic and mediastinal disorders | Pulmonary embolism | Unrelated | Severe | 22 | 32 | Recovered / Resolved |
| 101-018 | Worsening of Intracerebral Hemorrhage | Nervous system disorders | Cerebral haemorrhage | Unrelated | Severe | 0 | 6 | Fatal |
| 102-002 | Dural arteriovenous fistula | Nervous system disorders | Dural arteriovenous fistula | Unrelated | Moderate | 1 | 1 | Recovered / Resolved |
| 102-010 | Deep vein thrombosis in Left Popliteal Vein | Vascular disorders | Deep vein thrombosis | Unrelated | Severe | 75 | 83 | Recovered / Resolved |
| 103-001 | Respiratory distress | Respiratory, thoracic and mediastinal disorders | Respiratory distress | Unrelated | Moderate | 1 | 35 | Recovered / Resolved |
| 104-001 | Brain Herniation | Injury, poisoning and procedural complications | Brain herniation | Unrelated | Severe | 0 | 4 | Fatal |
| 106-002 | Focal Seizure | Nervous system disorders | Partial seizures | Unrelated | Severe | 8 | 18 | Recovered / Resolved |

**Supplemental Table 4.**
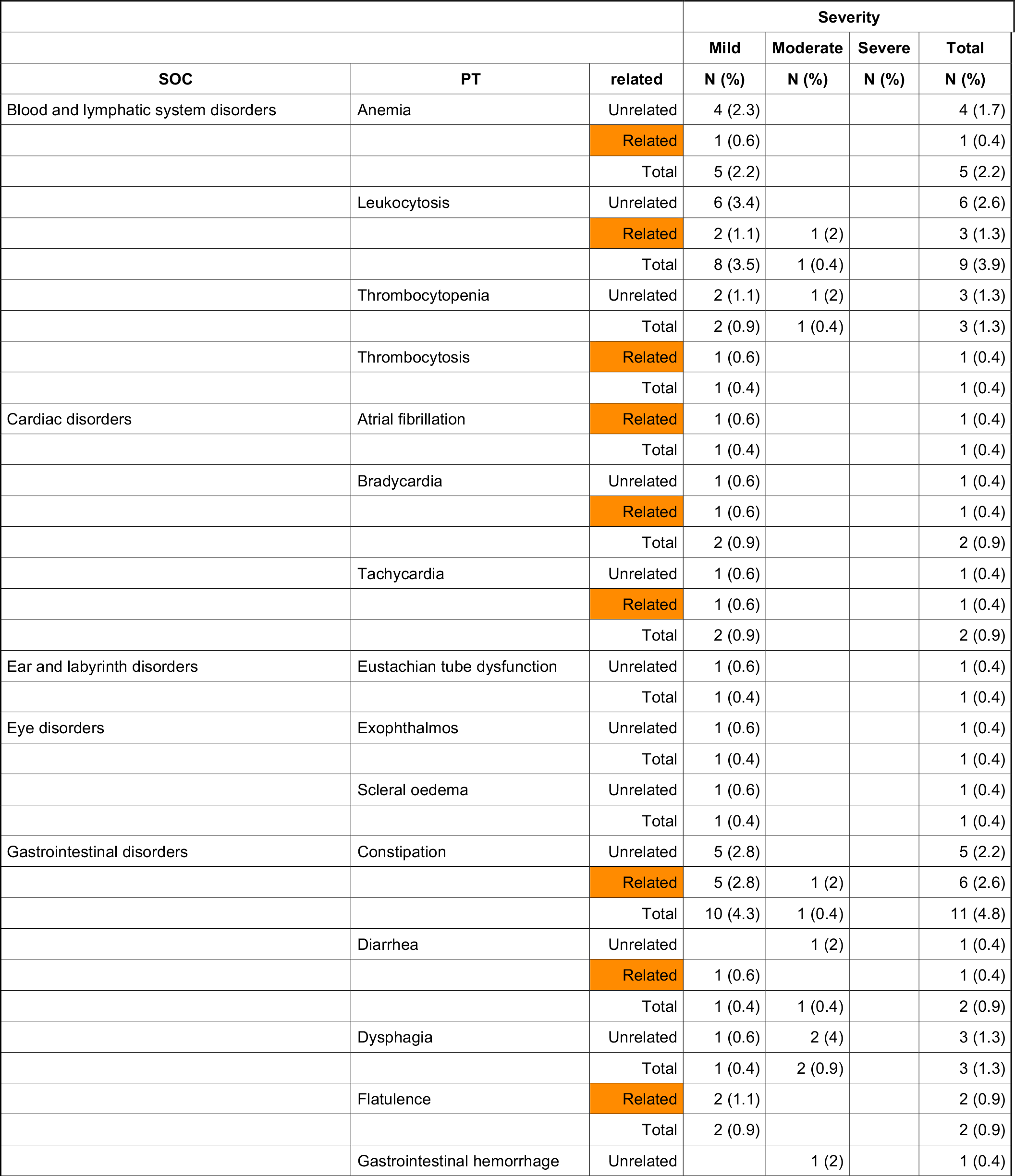
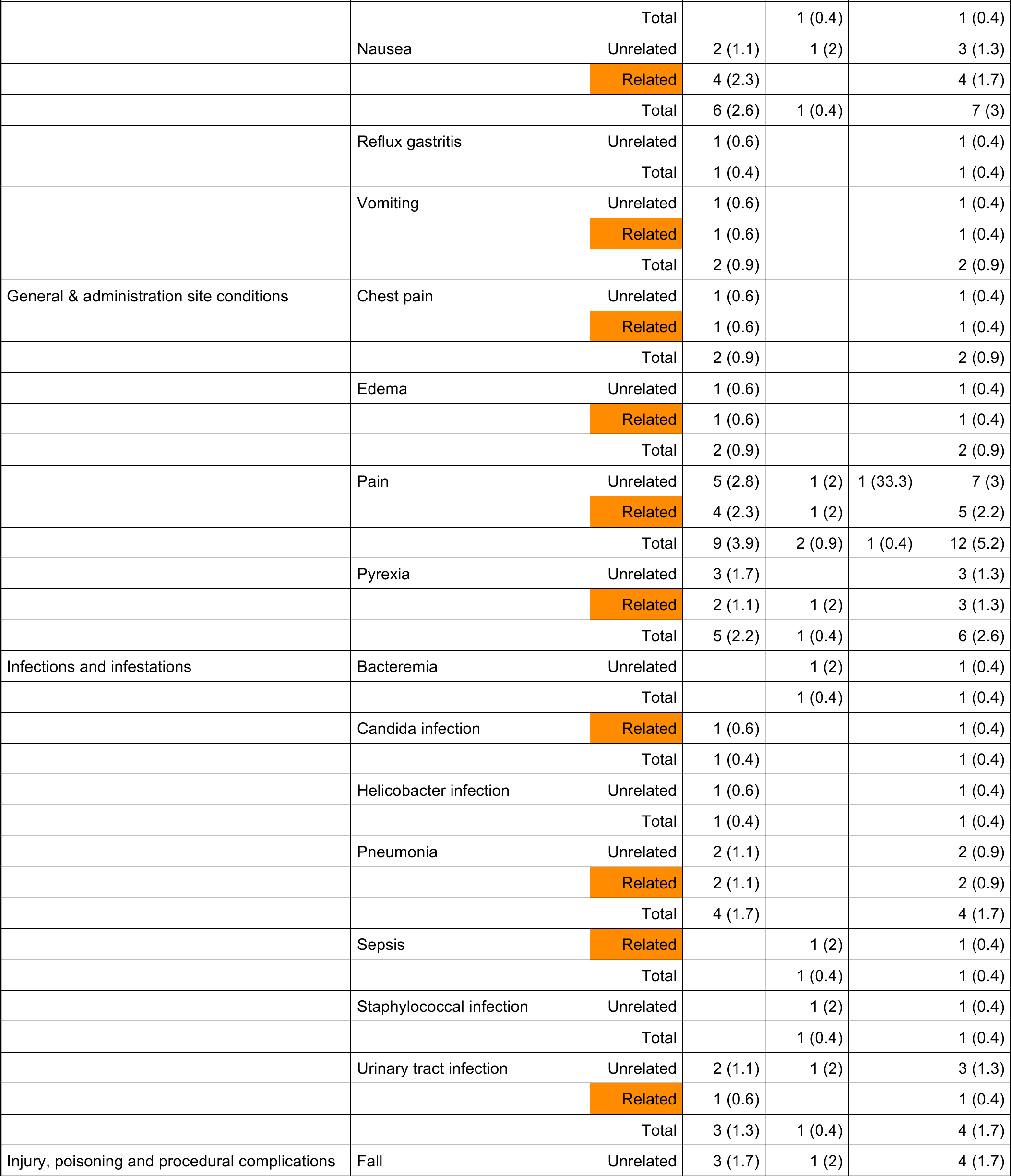
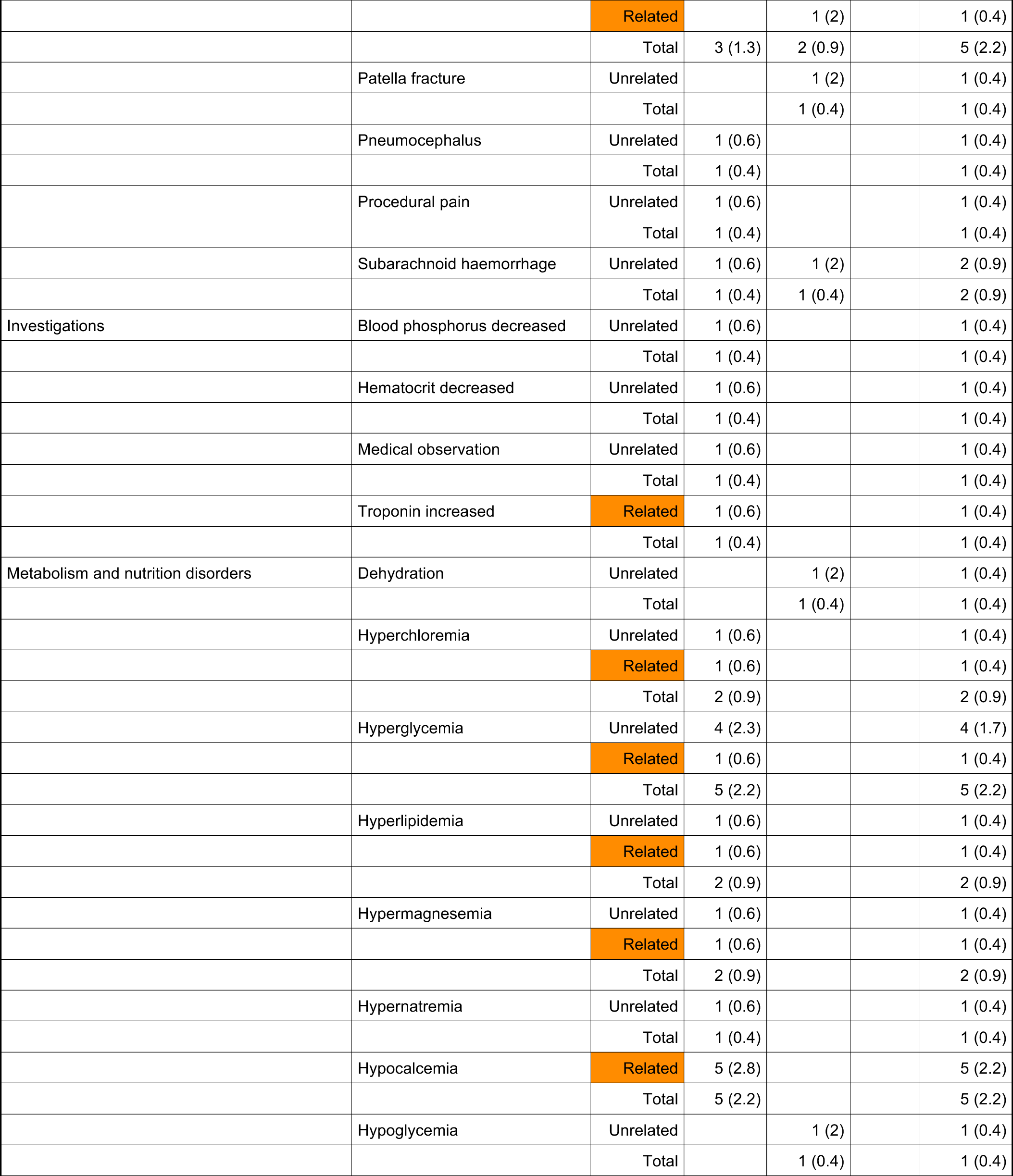
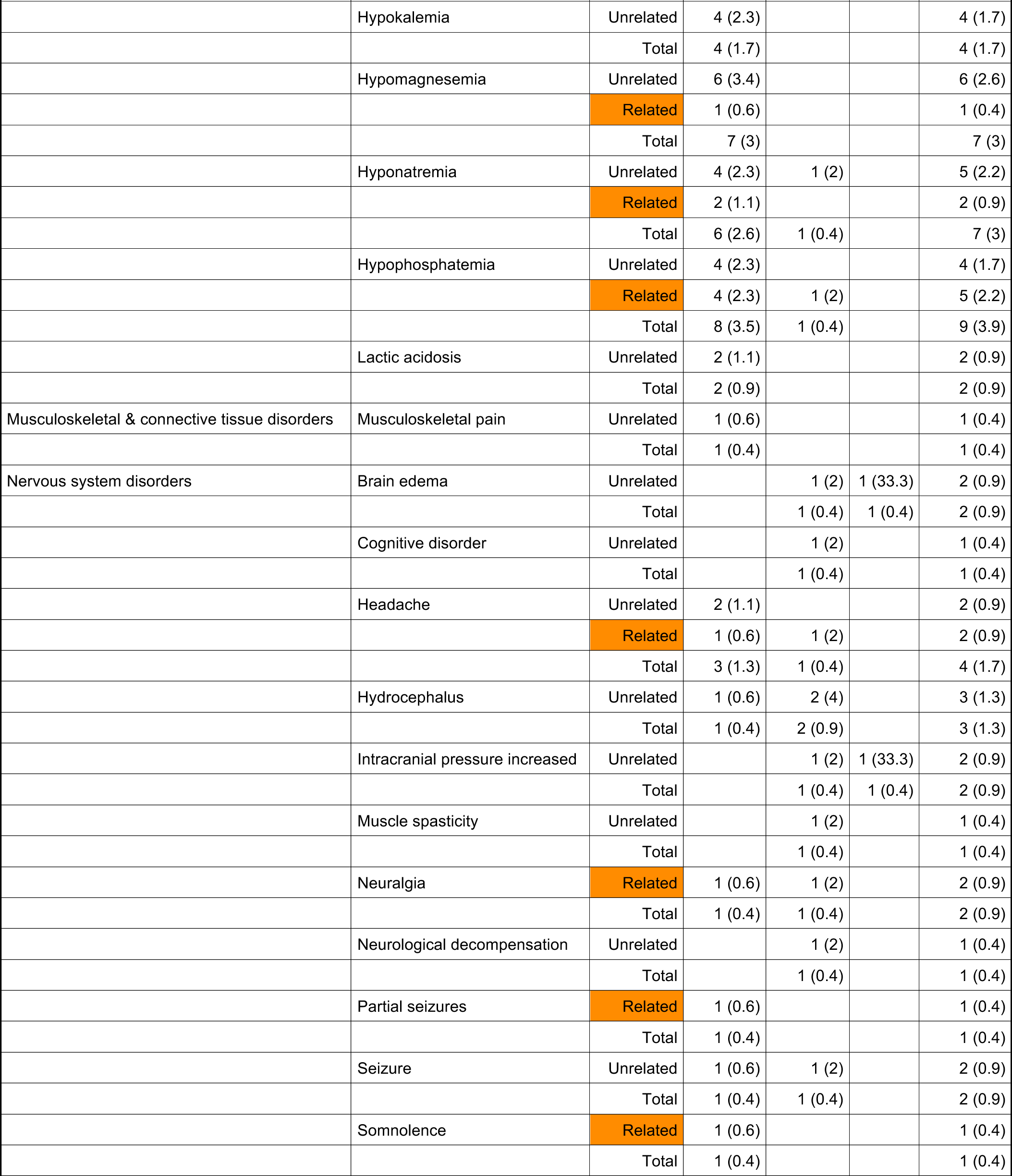
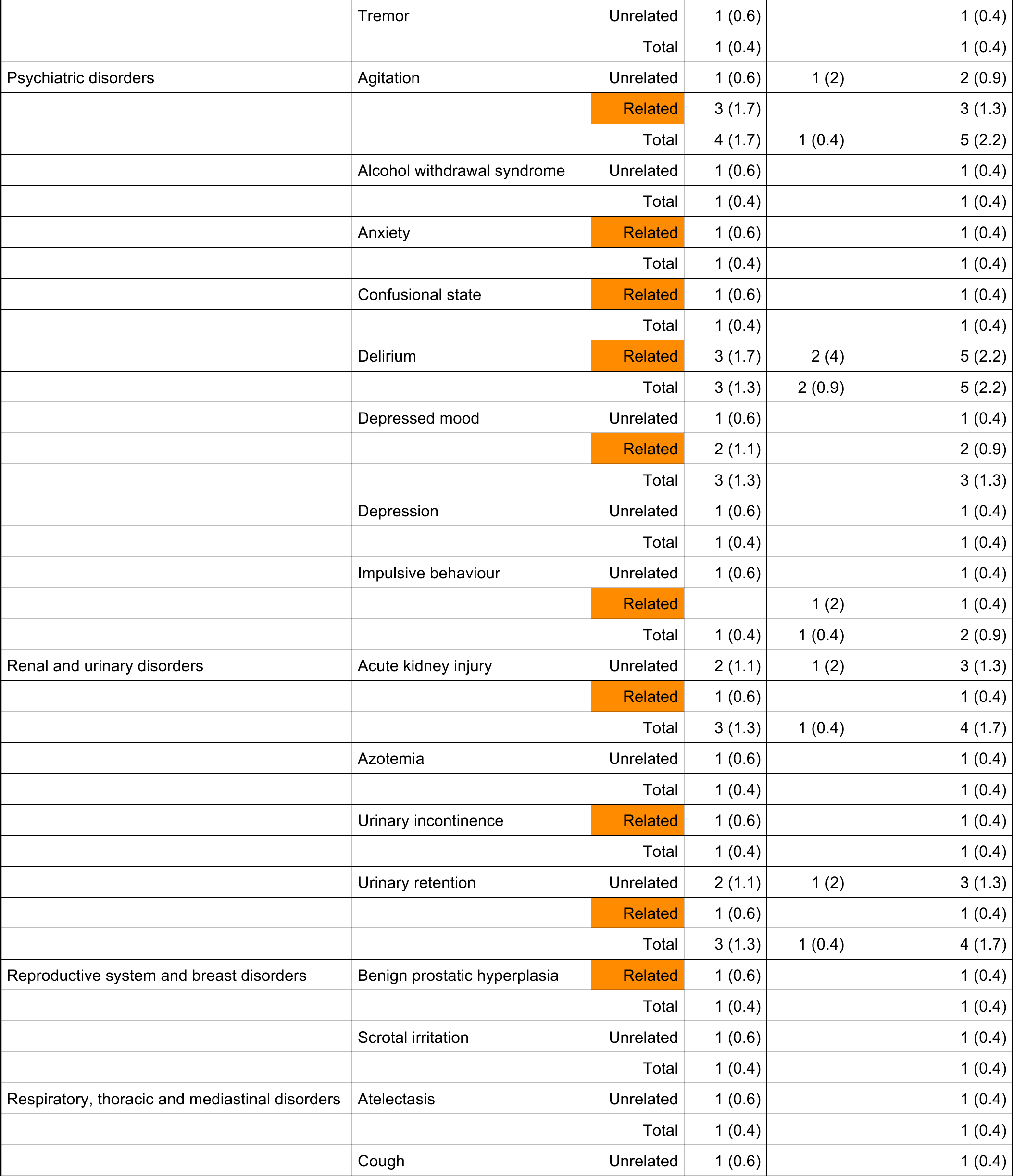
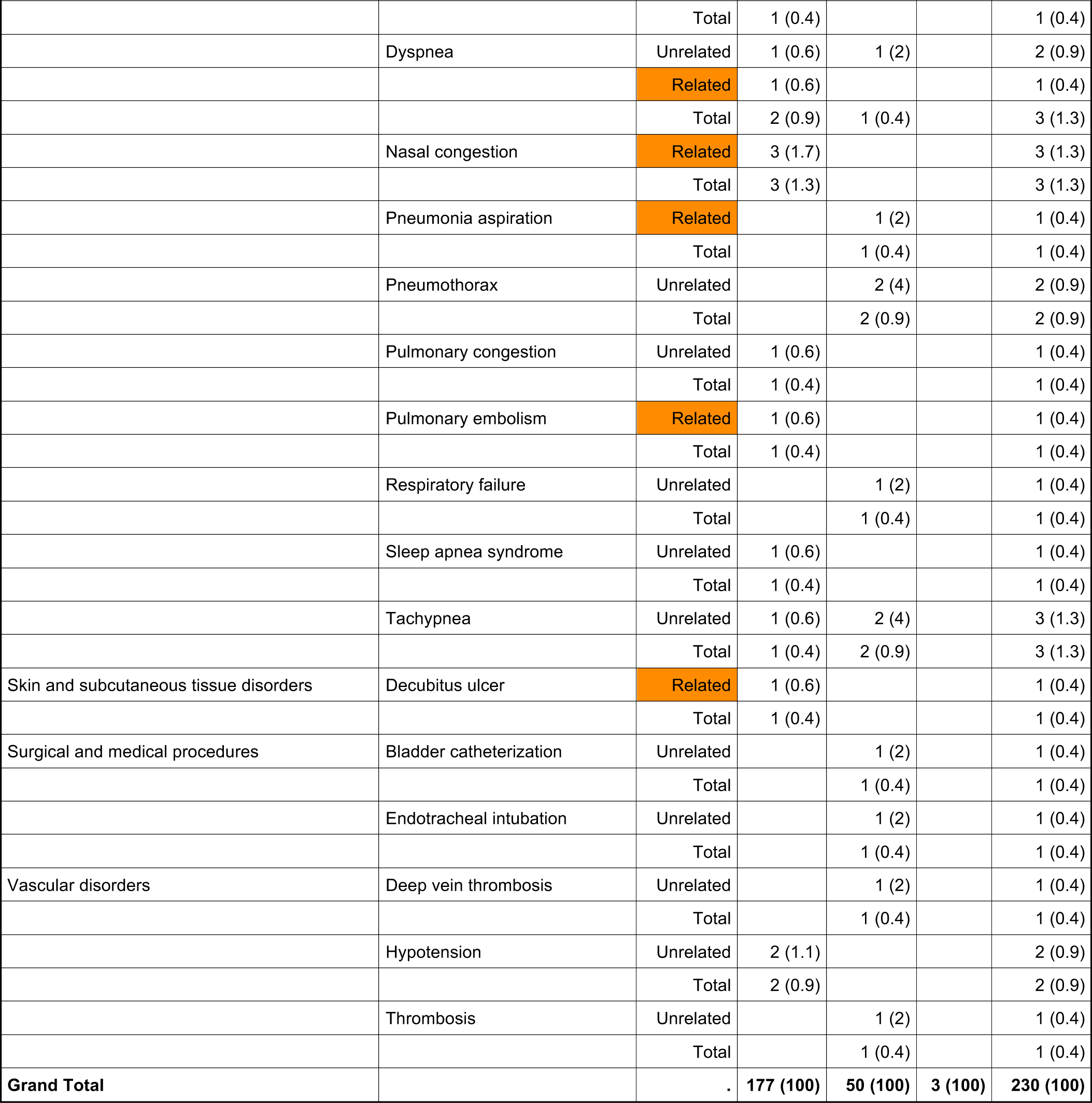
Full list of all non-serious treatment emergent adverse events in Participants with Acute SupraTentorial Intracerebral Hemorrhage (CATCH) Trial.

|  |  |  | Severity |  |  |  |
| --- | --- | --- | --- | --- | --- | --- |
|  |  |  | Mild | Moderate | Severe | Total |
| SOC | PT | related | N (%) | N (%) | N (%) | N (%) |
| Blood and lymphatic system disorders | Anemia | Unrelated | 4 (2.3) |  |  | 4 (1.7) |
|  |  | Related | 1 (0.6) |  |  | 1 (0.4) |
|  |  | Total | 5 (2.2) |  |  | 5 (2.2) |
|  | Leukocytosis | Unrelated | 6 (3.4) |  |  | 6 (2.6) |
|  |  | Related | 2 (1.1) | 1 (2) |  | 3 (1.3) |
|  |  | Total | 8 (3.5) | 1 (0.4) |  | 9 (3.9) |
|  | Thrombocytopenia | Unrelated | 2 (1.1) | 1 (2) |  | 3 (1.3) |
|  |  | Total | 2 (0.9) | 1 (0.4) |  | 3 (1.3) |
|  | Thrombocytosis | Related | 1 (0.6) |  |  | 1 (0.4) |
|  |  | Total | 1 (0.4) |  |  | 1 (0.4) |
| Cardiac disorders | Atrial fibrillation | Related | 1 (0.6) |  |  | 1 (0.4) |
|  |  | Total | 1 (0.4) |  |  | 1 (0.4) |
|  | Bradycardia | Unrelated | 1 (0.6) |  |  | 1 (0.4) |
|  |  | Related | 1 (0.6) |  |  | 1 (0.4) |
|  |  | Total | 2 (0.9) |  |  | 2 (0.9) |
|  | Tachycardia | Unrelated | 1 (0.6) |  |  | 1 (0.4) |
|  |  | Related | 1 (0.6) |  |  | 1 (0.4) |
|  |  | Total | 2 (0.9) |  |  | 2 (0.9) |
| Ear and labyrinth disorders | Eustachian tube dysfunction | Unrelated | 1 (0.6) |  |  | 1 (0.4) |
|  |  | Total | 1 (0.4) |  |  | 1 (0.4) |
| Eye disorders | Exophthalmos | Unrelated | 1 (0.6) |  |  | 1 (0.4) |
|  |  | Total | 1 (0.4) |  |  | 1 (0.4) |
|  | Scleral oedema | Unrelated | 1 (0.6) |  |  | 1 (0.4) |
|  |  | Total | 1 (0.4) |  |  | 1 (0.4) |
| Gastrointestinal disorders | Constipation | Unrelated | 5 (2.8) |  |  | 5 (2.2) |
|  |  | Related | 5 (2.8) | 1 (2) |  | 6 (2.6) |
|  |  | Total | 10 (4.3) | 1 (0.4) |  | 11 (4.8) |
|  | Diarrhea | Unrelated |  | 1 (2) |  | 1 (0.4) |
|  |  | Related | 1 (0.6) |  |  | 1 (0.4) |
|  |  | Total | 1 (0.4) | 1 (0.4) |  | 2 (0.9) |
|  | Dysphagia | Unrelated | 1 (0.6) | 2 (4) |  | 3 (1.3) |
|  |  | Total | 1 (0.4) | 2 (0.9) |  | 3 (1.3) |
|  | Flatulence | Related | 2 (1.1) |  |  | 2 (0.9) |
|  |  | Total | 2 (0.9) |  |  | 2 (0.9) |
|  | Gastrointestinal hemorrhage | Unrelated |  | 1 (2) |  | 1 (0.4) |
|  |  |  | Mild | Moderate | Severe | Total |
| SOC | PT | related | N (%) | N (%) | N (%) | N (%) |
|  |  | Total |  | 1 (0.4) |  | 1 (0.4) |
|  | Nausea | Unrelated | 2 (1.1) | 1 (2) |  | 3 (1.3) |
|  |  | Related | 4 (2.3) |  |  | 4 (1.7) |
|  |  | Total | 6 (2.6) | 1 (0.4) |  | 7 (3) |
|  | Reflux gastritis | Unrelated | 1 (0.6) |  |  | 1 (0.4) |
|  |  | Total | 1 (0.4) |  |  | 1 (0.4) |
|  | Vomiting | Unrelated | 1 (0.6) |  |  | 1 (0.4) |
|  |  | Related | 1 (0.6) |  |  | 1 (0.4) |
|  |  | Total | 2 (0.9) |  |  | 2 (0.9) |
| General & administration site conditions | Chest pain | Unrelated | 1 (0.6) |  |  | 1 (0.4) |
|  |  | Related | 1 (0.6) |  |  | 1 (0.4) |
|  |  | Total | 2 (0.9) |  |  | 2 (0.9) |
|  | Edema | Unrelated | 1 (0.6) |  |  | 1 (0.4) |
|  |  | Related | 1 (0.6) |  |  | 1 (0.4) |
|  |  | Total | 2 (0.9) |  |  | 2 (0.9) |
|  | Pain | Unrelated | 5 (2.8) | 1 (2) | 1 (33.3) | 7 (3) |
|  |  | Related | 4 (2.3) | 1 (2) |  | 5 (2.2) |
|  |  | Total | 9 (3.9) | 2 (0.9) | 1 (0.4) | 12 (5.2) |
|  | Pyrexia | Unrelated | 3 (1.7) |  |  | 3 (1.3) |
|  |  | Related | 2 (1.1) | 1 (2) |  | 3 (1.3) |
|  |  | Total | 5 (2.2) | 1 (0.4) |  | 6 (2.6) |
| Infections and infestations | Bacteremia | Unrelated |  | 1 (2) |  | 1 (0.4) |
|  |  | Total |  | 1 (0.4) |  | 1 (0.4) |
|  | Candida infection | Related | 1 (0.6) |  |  | 1 (0.4) |
|  |  | Total | 1 (0.4) |  |  | 1 (0.4) |
|  | Helicobacter infection | Unrelated | 1 (0.6) |  |  | 1 (0.4) |
|  |  | Total | 1 (0.4) |  |  | 1 (0.4) |
|  | Pneumonia | Unrelated | 2 (1.1) |  |  | 2 (0.9) |
|  |  | Related | 2 (1.1) |  |  | 2 (0.9) |
|  |  | Total | 4 (1.7) |  |  | 4 (1.7) |
|  | Sepsis | Related |  | 1 (2) |  | 1 (0.4) |
|  |  | Total |  | 1 (0.4) |  | 1 (0.4) |
|  | Staphylococcal infection | Unrelated |  | 1 (2) |  | 1 (0.4) |
|  |  | Total |  | 1 (0.4) |  | 1 (0.4) |
|  | Urinary tract infection | Unrelated | 2 (1.1) | 1 (2) |  | 3 (1.3) |
|  |  | Related | 1 (0.6) |  |  | 1 (0.4) |
|  |  | Total | 3 (1.3) | 1 (0.4) |  | 4 (1.7) |
| Injury, poisoning and procedural complications | Fall | Unrelated | 3 (1.7) | 1 (2) |  | 4 (1.7) |
|  |  |  | Mild | Moderate | Severe | Total |
| SOC | PT | related | N (%) | N (%) | N (%) | N (%) |
|  |  | Related |  | 1 (2) |  | 1 (0.4) |
|  |  | Total | 3 (1.3) | 2 (0.9) |  | 5 (2.2) |
|  | Patella fracture | Unrelated |  | 1 (2) |  | 1 (0.4) |
|  |  | Total |  | 1 (0.4) |  | 1 (0.4) |
|  | Pneumocephalus | Unrelated | 1 (0.6) |  |  | 1 (0.4) |
|  |  | Total | 1 (0.4) |  |  | 1 (0.4) |
|  | Procedural pain | Unrelated | 1 (0.6) |  |  | 1 (0.4) |
|  |  | Total | 1 (0.4) |  |  | 1 (0.4) |
|  | Subarachnoid haemorrhage | Unrelated | 1 (0.6) | 1 (2) |  | 2 (0.9) |
|  |  | Total | 1 (0.4) | 1 (0.4) |  | 2 (0.9) |
| Investigations | Blood phosphorus decreased | Unrelated | 1 (0.6) |  |  | 1 (0.4) |
|  |  | Total | 1 (0.4) |  |  | 1 (0.4) |
|  | Hematocrit decreased | Unrelated | 1 (0.6) |  |  | 1 (0.4) |
|  |  | Total | 1 (0.4) |  |  | 1 (0.4) |
|  | Medical observation | Unrelated | 1 (0.6) |  |  | 1 (0.4) |
|  |  | Total | 1 (0.4) |  |  | 1 (0.4) |
|  | Troponin increased | Related | 1 (0.6) |  |  | 1 (0.4) |
|  |  | Total | 1 (0.4) |  |  | 1 (0.4) |
| Metabolism and nutrition disorders | Dehydration | Unrelated |  | 1 (2) |  | 1 (0.4) |
|  |  | Total |  | 1 (0.4) |  | 1 (0.4) |
|  | Hyperchloremia | Unrelated | 1 (0.6) |  |  | 1 (0.4) |
|  |  | Related | 1 (0.6) |  |  | 1 (0.4) |
|  |  | Total | 2 (0.9) |  |  | 2 (0.9) |
|  | Hyperglycemia | Unrelated | 4 (2.3) |  |  | 4 (1.7) |
|  |  | Related | 1 (0.6) |  |  | 1 (0.4) |
|  |  | Total | 5 (2.2) |  |  | 5 (2.2) |
|  | Hyperlipidemia | Unrelated | 1 (0.6) |  |  | 1 (0.4) |
|  |  | Related | 1 (0.6) |  |  | 1 (0.4) |
|  |  | Total | 2 (0.9) |  |  | 2 (0.9) |
|  | Hypermagnesemia | Unrelated | 1 (0.6) |  |  | 1 (0.4) |
|  |  | Related | 1 (0.6) |  |  | 1 (0.4) |
|  |  | Total | 2 (0.9) |  |  | 2 (0.9) |
|  | Hypernatremia | Unrelated | 1 (0.6) |  |  | 1 (0.4) |
|  |  | Total | 1 (0.4) |  |  | 1 (0.4) |
|  | Hypocalcemia | Related | 5 (2.8) |  |  | 5 (2.2) |
|  |  | Total | 5 (2.2) |  |  | 5 (2.2) |
|  | Hypoglycemia | Unrelated |  | 1 (2) |  | 1 (0.4) |
|  |  | Total |  | 1 (0.4) |  | 1 (0.4) |
|  |  |  | Mild | Moderate | Severe | Total |
| SOC | PT | related | N (%) | N (%) | N (%) | N (%) |
|  | Hypokalemia | Unrelated | 4 (2.3) |  |  | 4 (1.7) |
|  |  | Total | 4 (1.7) |  |  | 4 (1.7) |
|  | Hypomagnesemia | Unrelated | 6 (3.4) |  |  | 6 (2.6) |
|  |  | Related | 1 (0.6) |  |  | 1 (0.4) |
|  |  | Total | 7 (3) |  |  | 7 (3) |
|  | Hyponatremia | Unrelated | 4 (2.3) | 1 (2) |  | 5 (2.2) |
|  |  | Related | 2 (1.1) |  |  | 2 (0.9) |
|  |  | Total | 6 (2.6) | 1 (0.4) |  | 7 (3) |
|  | Hypophosphatemia | Unrelated | 4 (2.3) |  |  | 4 (1.7) |
|  |  | Related | 4 (2.3) | 1 (2) |  | 5 (2.2) |
|  |  | Total | 8 (3.5) | 1 (0.4) |  | 9 (3.9) |
|  | Lactic acidosis | Unrelated | 2 (1.1) |  |  | 2 (0.9) |
|  |  | Total | 2 (0.9) |  |  | 2 (0.9) |
| Musculoskeletal & connective tissue disorders | Musculoskeletal pain | Unrelated | 1 (0.6) |  |  | 1 (0.4) |
|  |  | Total | 1 (0.4) |  |  | 1 (0.4) |
| Nervous system disorders | Brain edema | Unrelated |  | 1 (2) | 1 (33.3) | 2 (0.9) |
|  |  | Total |  | 1 (0.4) | 1 (0.4) | 2 (0.9) |
|  | Cognitive disorder | Unrelated |  | 1 (2) |  | 1 (0.4) |
|  |  | Total |  | 1 (0.4) |  | 1 (0.4) |
|  | Headache | Unrelated | 2 (1.1) |  |  | 2 (0.9) |
|  |  | Related | 1 (0.6) | 1 (2) |  | 2 (0.9) |
|  |  | Total | 3 (1.3) | 1 (0.4) |  | 4 (1.7) |
|  | Hydrocephalus | Unrelated | 1 (0.6) | 2 (4) |  | 3 (1.3) |
|  |  | Total | 1 (0.4) | 2 (0.9) |  | 3 (1.3) |
|  | Intracranial pressure increased | Unrelated |  | 1 (2) | 1 (33.3) | 2 (0.9) |
|  |  | Total |  | 1 (0.4) | 1 (0.4) | 2 (0.9) |
|  | Muscle spasticity | Unrelated |  | 1 (2) |  | 1 (0.4) |
|  |  | Total |  | 1 (0.4) |  | 1 (0.4) |
|  | Neuralgia | Related | 1 (0.6) | 1 (2) |  | 2 (0.9) |
|  |  | Total | 1 (0.4) | 1 (0.4) |  | 2 (0.9) |
|  | Neurological decompensation | Unrelated |  | 1 (2) |  | 1 (0.4) |
|  |  | Total |  | 1 (0.4) |  | 1 (0.4) |
|  | Partial seizures | Related | 1 (0.6) |  |  | 1 (0.4) |
|  |  | Total | 1 (0.4) |  |  | 1 (0.4) |
|  | Seizure | Unrelated | 1 (0.6) | 1 (2) |  | 2 (0.9) |
|  |  | Total | 1 (0.4) | 1 (0.4) |  | 2 (0.9) |
|  | Somnolence | Related | 1 (0.6) |  |  | 1 (0.4) |
|  |  | Total | 1 (0.4) |  |  | 1 (0.4) |
|  |  |  | Mild | Moderate | Severe | Total |
| SOC | PT | related | N (%) | N (%) | N (%) | N (%) |
|  | Tremor | Unrelated | 1 (0.6) |  |  | 1 (0.4) |
|  |  | Total | 1 (0.4) |  |  | 1 (0.4) |
| Psychiatric disorders | Agitation | Unrelated | 1 (0.6) | 1 (2) |  | 2 (0.9) |
|  |  | Related | 3 (1.7) |  |  | 3 (1.3) |
|  |  | Total | 4 (1.7) | 1 (0.4) |  | 5 (2.2) |
|  | Alcohol withdrawal syndrome | Unrelated | 1 (0.6) |  |  | 1 (0.4) |
|  |  | Total | 1 (0.4) |  |  | 1 (0.4) |
|  | Anxiety | Related | 1 (0.6) |  |  | 1 (0.4) |
|  |  | Total | 1 (0.4) |  |  | 1 (0.4) |
|  | Confusional state | Related | 1 (0.6) |  |  | 1 (0.4) |
|  |  | Total | 1 (0.4) |  |  | 1 (0.4) |
|  | Delirium | Related | 3 (1.7) | 2 (4) |  | 5 (2.2) |
|  |  | Total | 3 (1.3) | 2 (0.9) |  | 5 (2.2) |
|  | Depressed mood | Unrelated | 1 (0.6) |  |  | 1 (0.4) |
|  |  | Related | 2 (1.1) |  |  | 2 (0.9) |
|  |  | Total | 3 (1.3) |  |  | 3 (1.3) |
|  | Depression | Unrelated | 1 (0.6) |  |  | 1 (0.4) |
|  |  | Total | 1 (0.4) |  |  | 1 (0.4) |
|  | Impulsive behaviour | Unrelated | 1 (0.6) |  |  | 1 (0.4) |
|  |  | Related |  | 1 (2) |  | 1 (0.4) |
|  |  | Total | 1 (0.4) | 1 (0.4) |  | 2 (0.9) |
| Renal and urinary disorders | Acute kidney injury | Unrelated | 2 (1.1) | 1 (2) |  | 3 (1.3) |
|  |  | Related | 1 (0.6) |  |  | 1 (0.4) |
|  |  | Total | 3 (1.3) | 1 (0.4) |  | 4 (1.7) |
|  | Azotemia | Unrelated | 1 (0.6) |  |  | 1 (0.4) |
|  |  | Total | 1 (0.4) |  |  | 1 (0.4) |
|  | Urinary incontinence | Related | 1 (0.6) |  |  | 1 (0.4) |
|  |  | Total | 1 (0.4) |  |  | 1 (0.4) |
|  | Urinary retention | Unrelated | 2 (1.1) | 1 (2) |  | 3 (1.3) |
|  |  | Related | 1 (0.6) |  |  | 1 (0.4) |
|  |  | Total | 3 (1.3) | 1 (0.4) |  | 4 (1.7) |
| Reproductive system and breast disorders | Benign prostatic hyperplasia | Related | 1 (0.6) |  |  | 1 (0.4) |
|  |  | Total | 1 (0.4) |  |  | 1 (0.4) |
|  | Scrotal irritation | Unrelated | 1 (0.6) |  |  | 1 (0.4) |
|  |  | Total | 1 (0.4) |  |  | 1 (0.4) |
| Respiratory, thoracic and mediastinal disorders | Atelectasis | Unrelated | 1 (0.6) |  |  | 1 (0.4) |
|  |  | Total | 1 (0.4) |  |  | 1 (0.4) |
|  | Cough | Unrelated | 1 (0.6) |  |  | 1 (0.4) |
|  |  |  | Mild | Moderate | Severe | Total |
| SOC | PT | related | N (%) | N (%) | N (%) | N (%) |
|  |  | Total | 1 (0.4) |  |  | 1 (0.4) |
|  | Dyspnea | Unrelated | 1 (0.6) | 1 (2) |  | 2 (0.9) |
|  |  | Related | 1 (0.6) |  |  | 1 (0.4) |
|  |  | Total | 2 (0.9) | 1 (0.4) |  | 3 (1.3) |
|  | Nasal congestion | Related | 3 (1.7) |  |  | 3 (1.3) |
|  |  | Total | 3 (1.3) |  |  | 3 (1.3) |
|  | Pneumonia aspiration | Related |  | 1 (2) |  | 1 (0.4) |
|  |  | Total |  | 1 (0.4) |  | 1 (0.4) |
|  | Pneumothorax | Unrelated |  | 2 (4) |  | 2 (0.9) |
|  |  | Total |  | 2 (0.9) |  | 2 (0.9) |
|  | Pulmonary congestion | Unrelated | 1 (0.6) |  |  | 1 (0.4) |
|  |  | Total | 1 (0.4) |  |  | 1 (0.4) |
|  | Pulmonary embolism | Related | 1 (0.6) |  |  | 1 (0.4) |
|  |  | Total | 1 (0.4) |  |  | 1 (0.4) |
|  | Respiratory failure | Unrelated |  | 1 (2) |  | 1 (0.4) |
|  |  | Total |  | 1 (0.4) |  | 1 (0.4) |
|  | Sleep apnea syndrome | Unrelated | 1 (0.6) |  |  | 1 (0.4) |
|  |  | Total | 1 (0.4) |  |  | 1 (0.4) |
|  | Tachypnea | Unrelated | 1 (0.6) | 2 (4) |  | 3 (1.3) |
|  |  | Total | 1 (0.4) | 2 (0.9) |  | 3 (1.3) |
| Skin and subcutaneous tissue disorders | Decubitus ulcer | Related | 1 (0.6) |  |  | 1 (0.4) |
|  |  | Total | 1 (0.4) |  |  | 1 (0.4) |
| Surgical and medical procedures | Bladder catheterization | Unrelated |  | 1 (2) |  | 1 (0.4) |
|  |  | Total |  | 1 (0.4) |  | 1 (0.4) |
|  | Endotracheal intubation | Unrelated |  | 1 (2) |  | 1 (0.4) |
|  |  | Total |  | 1 (0.4) |  | 1 (0.4) |
| Vascular disorders | Deep vein thrombosis | Unrelated |  | 1 (2) |  | 1 (0.4) |
|  |  | Total |  | 1 (0.4) |  | 1 (0.4) |
|  | Hypotension | Unrelated | 2 (1.1) |  |  | 2 (0.9) |
|  |  | Total | 2 (0.9) |  |  | 2 (0.9) |
|  | Thrombosis | Unrelated |  | 1 (2) |  | 1 (0.4) |
|  |  | Total |  | 1 (0.4) |  | 1 (0.4) |
| **Grand Total** |  | . | **177 (100)** | **50 (100)** | **3 (100)** | **230 (100)** |

**Supplemental Table 5.**
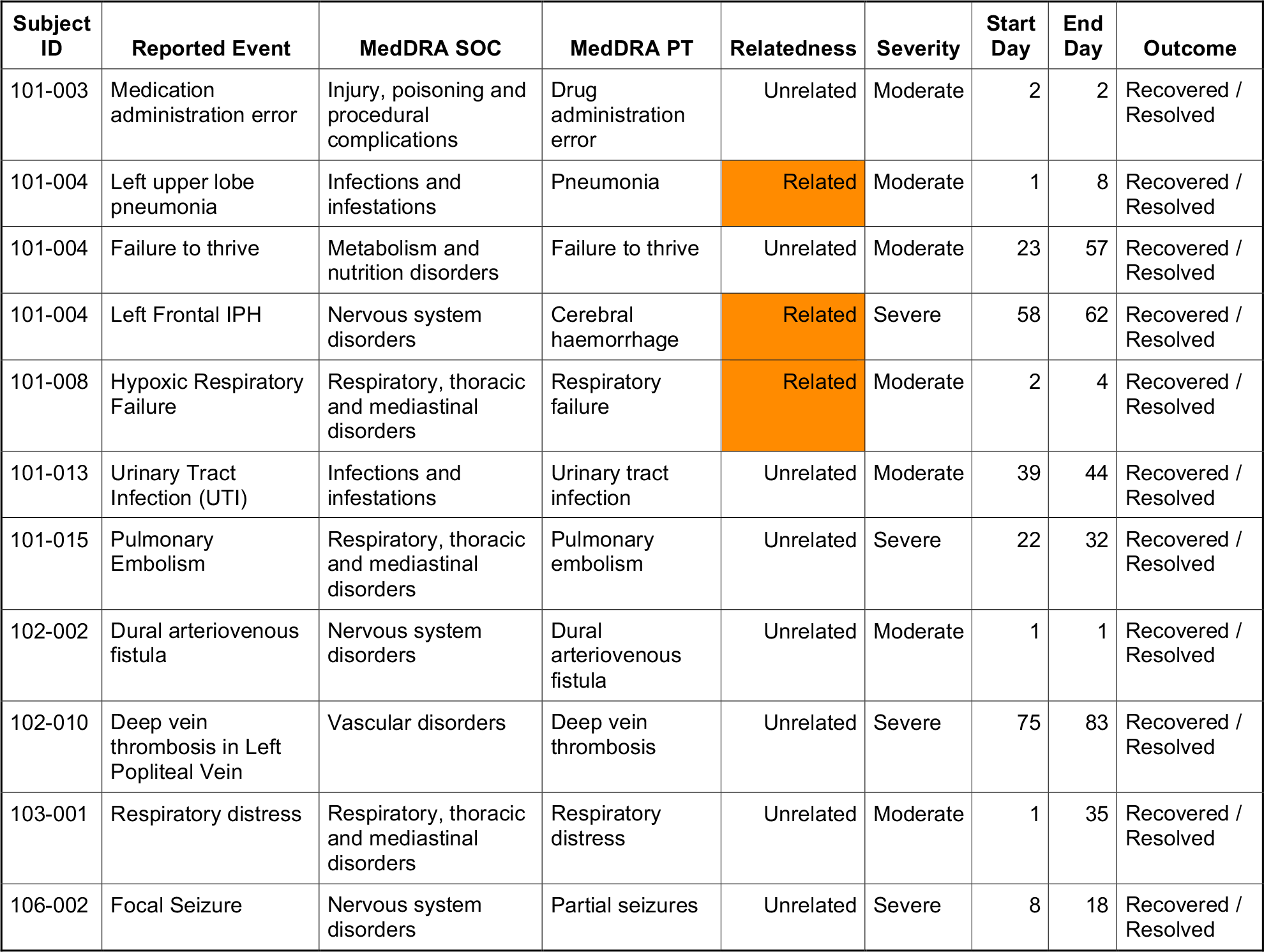
Full list of all treatment emergent serious adverse events in Participants with Acute SupraTentorial Intracerebral Hemorrhage (CATCH) Trial. Participants with Acute SupraTentorial Intracerebral Hemorrhage (CATCH) Trial. Participants with Acute SupraTentorial Intracerebral Hemorrhage (CATCH) Trial. Participants with Acute SupraTentorial Intracerebral Hemorrhage (CATCH) Trial. Barthel Index: The median (range) of the change (Day 30 minus Day 5, N=28) was 15 (−30, 75) with mean (SD) of 17.14 (25.18) (P=0.001, Wilcoxon signed rank test). MOCA: The median (range) of the change (Day 30 minus Day 5, N=17) was 1 (−4, 10) with mean (SD) of 2.47 (4.33) (P=0.04, Wilcoxon signed rank test). SIS-16: The median (range) of the change (Day 30 minus Day 5, N=24) was 10 (−48, 39) with mean (SD) of 8.63 (18.43) (P=0.01, Wilcoxon signed rank test). NIHSS: P-for-trend = 0.01 over time from Day 0 (Enrollment and First Dose) through Day 30 based on a normal generalized estimating equation with unstructured working correlation.

| Subject ID | Reported Event | MedDRA SOC | MedDRA PT | Relatedness | Severity | Start Day | End Day | Outcome |
| --- | --- | --- | --- | --- | --- | --- | --- | --- |
| 101-003 | Medication administration error | Injury, poisoning and procedural complications | Drug administration error | Unrelated | Moderate | 2 | 2 | Recovered / Resolved |
| 101-004 | Left upper lobe pneumonia | Infections and infestations | Pneumonia | Related | Moderate | 1 | 8 | Recovered / Resolved |
| 101-004 | Failure to thrive | Metabolism and nutrition disorders | Failure to thrive | Unrelated | Moderate | 23 | 57 | Recovered / Resolved |
| 101-004 | Left Frontal IPH | Nervous system disorders | Cerebral haemorrhage | Related | Severe | 58 | 62 | Recovered / Resolved |
| 101-008 | Hypoxic Respiratory Failure | Respiratory, thoracic and mediastinal disorders | Respiratory failure | Related | Moderate | 2 | 4 | Recovered / Resolved |
| 101-013 | Urinary Tract Infection (UTI) | Infections and infestations | Urinary tract infection | Unrelated | Moderate | 39 | 44 | Recovered / Resolved |
| 101-015 | Pulmonary Embolism | Respiratory, thoracic and mediastinal disorders | Pulmonary embolism | Unrelated | Severe | 22 | 32 | Recovered / Resolved |
| 102-002 | Dural arteriovenous fistula | Nervous system disorders | Dural arteriovenous fistula | Unrelated | Moderate | 1 | 1 | Recovered / Resolved |
| 102-010 | Deep vein thrombosis in Left Popliteal Vein | Vascular disorders | Deep vein thrombosis | Unrelated | Severe | 75 | 83 | Recovered / Resolved |
| 103-001 | Respiratory distress | Respiratory, thoracic and mediastinal disorders | Respiratory distress | Unrelated | Moderate | 1 | 35 | Recovered / Resolved |
| 106-002 | Focal Seizure | Nervous system disorders | Partial seizures | Unrelated | Severe | 8 | 18 | Recovered / Resolved |

**Supplemental Table 6.** CATCH Secondary Outcomes.

|  | Screening<br>Visit 1, Day<br>0 | Visit 2, Day 0 –<br>Enrollment<br>and First Dose | Visit 3, Day<br>1 -<br>Treatment | Visit 4, Day<br>2 -<br>Treatment | Visit 5, Day<br>3 -<br>Treatment | Visit 6, Day<br>4 -<br>Treatment | Visit 7, Day<br>5 -<br>Treatment | Visit 8,<br>Day 30 -<br>30 Day<br>Follow-up |
| --- | --- | --- | --- | --- | --- | --- | --- | --- |
| **Barthel**<br>**Index** |  |  |  |  |  |  |  |  |
| N |  |  |  |  |  |  | 32 | 30 |
| Mean<br>(SD) |  |  |  |  |  |  | 38.3 (31.8) | 55.8 (37.6) |
| Median<br>(Min,<br>Max) |  |  |  |  |  |  | 32.5 (0.0,<br>100.0) | 47.5 (0.0,<br>100.0) |
| **MOCA** |  |  |  |  |  |  |  |  |
| N |  |  |  |  |  |  | 27 | 20 |
| Mean<br>(SD) |  |  |  |  |  |  | 15.4 (10.7) | 19.1 (9.6) |
| Median<br>(Min,<br>Max) |  |  |  |  |  |  | 19.0 (0.0,<br>30.0) | 22.0 (0.0,<br>30.0) |
| **NIHSS** |  |  |  |  |  |  |  |  |
| N | 38 | 38 | 38 | 36 | 36 | 34 | 34 | 23 |
| Mean<br>(SD) | 13.0 (7.4) | 13.1 (6.7) | 13.0 (7.5) | 11.7 (7.7) | 11.4 (8.5) | 11.8 (8.4) | 11.1 (8.3) | 6.9 (6.1) |
| Median<br>(Min,<br>Max) | 10.5 (4.0,<br>29.0) | 12.0 (4.0, 25.0) | 11.0 (1.0,<br>29.0) | 9.5 (2.0,<br>29.0) | 8.0 (1.0,<br>36.0) | 9.0 (1.0,<br>31.0) | 8.5 (1.0,<br>29.0) | 5.0 (0.0,<br>21.0) |
| **SIS-16** |  |  |  |  |  |  |  |  |
| N |  |  |  |  |  |  | 27 | 27 |
| Mean<br>(SD) |  |  |  |  |  |  | 37.2 (21.1) | 45.4 (22.7) |
| Median<br>(Min,<br>Max) |  |  |  |  |  |  | 30.0 (16.0,<br>79.0) | 43.0 (16.0,<br>79.0) |

**Supplemental Figure.**
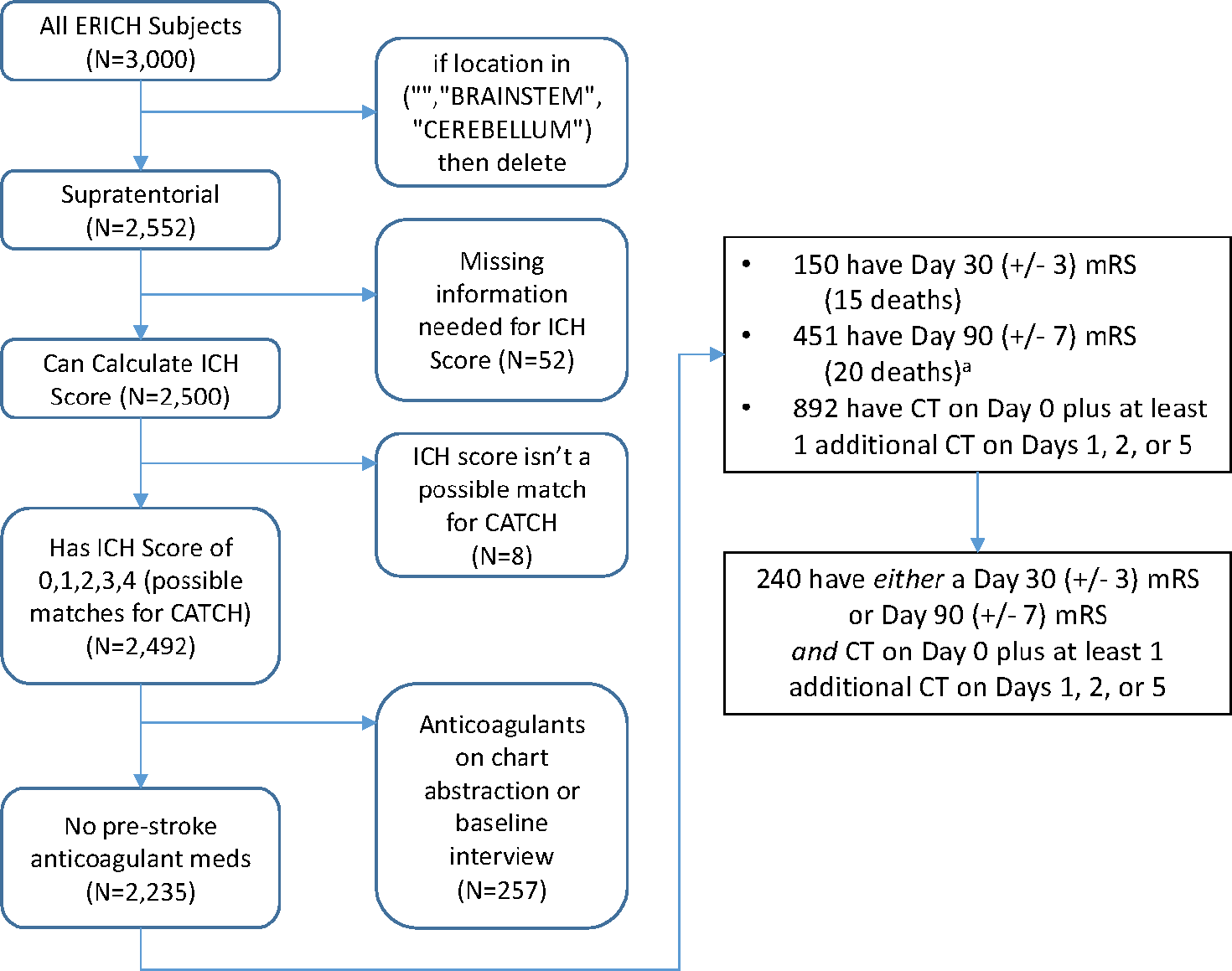
Consort diagram for ERICH cohort matching to CATCH trial participants. Consort diagram for matching Ethnic/Racial Variations of Intracerebral Hemorrhage (ERICH) study cohort to CN-105 in Participants with Acute SupraTentorial Intracerebral Hemorrhage (CATCH) Trial participants. ERICH participants were matched 1:1 to CATCH participants based on initial ICH Score, supratentorial location, absence of anticoagulation, and demographic variables including age, sex, race/ethnicity, history of diabetes and hypertension.

